# Investigating rehabilitation by activities involving the trunk to improve balance and gait control in young children with cerebral palsy: a randomized open-label crossover trial protocol

**DOI:** 10.1101/2025.10.02.25336667

**Authors:** Stella Zografou, Jonathan Pierret, Rajul Vasa, Jean Paysant, Christian Beyaert

## Abstract

**Introduction:** Children with cerebral palsy (CP) have gross motor and balance disorders altering standing, walking and activities. Since trunk control is central for balance, rehabilitation targeting the trunk is developing. In children with CP aged 5 to 12 years, rehabilitation by activities involving the trunk (RAIT) based on activities in intermediate postures for 3 months has been demonstrated to significantly improve trunk control while standing and early trunk deceleration and coupled negative ankle power due to plantar flexors while walking autonomously. As motor disorders develop early, the effects of RAIT are investigated in younger children and for longer time: the adapted design of this study is presented. Initial motor disorders in children with CP aged 18 months to 5 years and 6 months, compared with typically developing children, are expected to be better reduced after RAIT during its first 3 months application than after usual rehabilitation, and to be increasingly reduced after 3, 6 and 12 months of RAIT.

**Methods:** The studied motor disorders include −1-during gait, excessive early anterior deceleration of the sternum (primary outcome) measured by inertial measurement unit, excessive anterior location of center of pressure on affected leg(s), excessive enhanced gait variability index and step width measured by a walkway equipped with pressure sensors, −2-Altered gross motor, balance and trunk function measured by the item set version of the Gross Motor Function Measurement 66 and by the Early Clinical Assessment of Balance.

**Expected results:** All these variables would be influenced by trunk balance and control, and therefore reduced after RAIT.

**Trial registration:** ClinicalTrials.gov **ID: NCT06438432**

**What this paper adds?:**

- Balance control is essential in motor control with a central role of the trunk.
- Children with cerebral palsy (CP) have motor, balance and trunk deficits.
- A trunk-focused rehabilitation by activities in intermediate postures presented (RAIT).
- Study design to assess the long-term effects of RAIT in young children with CP.
- RAIT would improve trunk and foot dynamics during gait and gross motor function.

## 1. Introduction

Cerebral Palsy (CP) is the most frequent disability of childhood that disturbs motor function with a prevalence of 2-3 births in every 1000 births (1). Cerebral palsy is a group of permanent but not unchanging disorders of posture and movement which are attributed to non-progressive brain damage occurring during perinatal development (2,3). The International Classification of Functioning, Disability and Health - children and youth version (ICF-CY) is a good way of addressing the impact of these deficits in children with CP. The ICF-CY provides a conceptual and systematic framework based on a biopsychosocial approach to standardize the health and health-related states of various pathological populations. Disability and function are described through the following three main domains: body functions and structures, activity and participation (4). Cerebral palsy affects these three levels, with structural alterations of the neural and musculoskeletal systems (5) affecting posture, activity and participation (6).

Indeed, postural control ensure safety balance against gravity and regulate the orientation and the position of body segments relative to the environment (7,8) in order to interact with the latter during voluntary movement. Its development relies on the proper maturation of the central nervous system (9), which is influenced by internal, such as the maturation of sensory information processing, and environmental constraints. These processes take place from the first year of life and continue during early childhood. The effective development of postural control is closely linked to the emergence of gross motor skills, which influences the field of activity and participation (10). In fact, all interaction with and within the environment involves a postural component and others.

The axial segments, in particular the trunk, play a key role in the development of postural control, with an impact on the different domains of the ICF. When trunk control is sufficiently developed, it provides a stable sitting posture that allows for the development of gross motor function (11), manual skills (12,13) and interactions with others (14). Later on, during childhood, spontaneous postural oscillations when standing decrease as the child grows. This decrease in oscillations is associated to improvement in postural control (15), where proactive control completes reactive control of disequilibrium (16), with an important part of trunk control (17).

Independent standing, which requires effective tonic antigravity activity of the trunk muscles, is an essential prerequisite for the onset of autonomous walking. The organization of the locomotor pattern, which emerges between 11 and 15 months (18,19), depends in large part on the progressive mastery of the coordination of axial segments during childhood (20–22). The importance of trunk control during walking is present even in adulthood, contributing not only to the maintenance of locomotor balance but also to propulsion (23–27).

From infancy onwards, children with CP show impaired control of axial segments and particularly of the trunk. These disorders appear first in the sitting position with abnormal postural reaction compared to children with typical development (TD) (28) and persist throughout childhood with combined impairment of static and dynamic control, with probable disturbance of proactive control mechanisms (29–31). The correlation between impaired trunk control and deficits in gross motor function has also been confirmed in children with CP (11).

Independent walking is a motor function that is crucial for autonomy, enabling the majority of activities of daily life to be carried out within the community. During walking, children with CP show deviations in head and trunk kinematics and kinetics in all three planes compared to children with TD (32–35). These deviations are associated with altered balance control during walking (36), leading to greater step width and variability in step length (37,38) and increased accelerations of the axial segments in all three planes (39,40). The latter is significantly related to trunk control deficit (41,42) that is the primary contributor to impaired walking performance in children with cerebral palsy, above neuromuscular deficits (43). Since the trunk and lower limbs interact reciprocally during walking, deviations of the trunk can induce deviations of the lower limbs, and vice versa (41).

Toe walking, characterized by the absence of the first heel pivot from initial contact (44), is common in children with CP. During weight acceptance (WA) phase of gait (defined as the period of combined initial power absorption activity around the lower joints (45), toe walking is associated with an early ankle power absorption and negative work, resulting in early decelerated ankle dorsiflexion and anterior tibial tilt (46). While the spastic origin of toe walking, i.e. related to hyperexcitable stretch reflexes, has been debated for decades and is now considered unlikely (47), authors have proposed this walking pattern to be part of an adaptive process (46,48). Given that the role of plantar flexors during WA is mainly to control trunk and body support and balance through their action on the tibia as soon as the foot is flat on the floor (49–51) toe walking allows the plantar flexors to early control the trunk from initial contact, suggesting their potential role to compensate for trunk control deficits that are associated to trunk lower stability (42). In a recent report from our research team (52), this adaptive role of the plantar flexors has been supported −1-by the significant correlation between negative ankle power and both the anterior deceleration of the upper trunk and the downward deceleration of the sacrum during WA; and −2-the simultaneous reduction in the upper trunk and sacrum deceleration and the negative ankle work during WA following trunk-focused rehabilitation resulting in significant improvement in trunk control.

Considering the significant role of the trunk in static and dynamic activities, as well as its impact on gross motor function from early childhood, rehabilitation targeting the trunk is being explored in children with CP (53–55) but it’s long term outcomes remains unexplored. In children with CP aged 5 to 12 years walking autonomously, trunk-focused rehabilitation (RAIT) but not usual rehabilitation (UR), for 3 months each, significantly improved trunk control while sitting and standing and early trunk deceleration and coupled negative ankle power due to plantar flexors (52). As these findings are promising, and since trunk control deficits appear early in children with CP, it is worth studying the effects of RAIT applied in younger children and for longer time. However, the protocol of this study needs specific adaptations in the content of RAIT and in technical aspects of assessing trunk control, gross motor control and gait dynamics. The aims and protocol of this project are presented and discussed below.

The aims of this randomized open-label crossover trial are to evaluate the motor effects of RAIT along its 9 to 12 months application and comparatively to usual rehabilitation (UR) during its first 3 months application in young children with CP who walk independently or with inconsistent use of a walking aid. Thus, initial motor disorders in children with CP aged 18 months to 5 years and 6 months, compared with children with TD, are expected to be better reduced after RAIT than after UR, and to be increasingly reduced after 3, 6 and 12 months of RAIT. Motor evaluation is carried out with consideration for the young age of the children. Trunk analysis during gait and gait analysis are based on inertial measurement units (IMUs) placed on the trunk for upper and lower trunk acceleration, on the use of a walkway equipped with pressure sensors for temporo-spatial parameters and center of pressure of the feet, and video recording for the Edinburgh visual gait score (56). The Item Set version of the Gross Motor Function Measurement 66 (GMFM-66-IS) (57) and the Early Clinical Assessment of Balance (ECAB) (58) are used to assess gross motor function and early balance and trunk function, respectively.

Initial motor disorders in young children with CP compared to children with TD are hypothesized to affect: −1-balance and trunk control and gross motor function: decrease in ECAB and GMFM-66-IS scores; −2-trunk dynamics during WA of gait: increase in anterior deceleration of the sternum (primary outcome) and downward deceleration of the waist at the level of the L5 vertebra (closed to body center of mass); −3-gait pattern related to balance disorder: increase in the enhanced gait variability index (eGVI) and in step width; −4-gait pattern including a toe walking: excessive anterior location of center of pressure (CoP) during WA and decrease in the Edinburgh Visual Gait Score.

As balance disorders with a primary role of trunk control deficit are believed to be essential factors of these motor disorders in children with CP, we hypothesize a significant correlation between all these variables. For the same reason, all these variables are expected to better improve after RAIT than after UR, and to increasingly improve after 3, 6 and 12 months of RAIT.

In addition, as upper limb(s) function may be affected in children with CP with consequences in child’s activity and participation, the parents are asked to complete the Reach Out Questionnaire that allows a holistic overview of functioning including activity limitations and participation (59). In our experience in children with hemiplegic and triplegic CP, RAIT improved the functional use of hand and upper limb and the child’s participation in various activities involving the upper limb. Thus, the Reach Out Questionnaire score is hypothesized to be reduced in children with CP compared to children with TD and to be increased more significantly after RAIT than after UR and further increased as RAIT is prolonged.

## 2. Materials and methods

### 2.1 Ethics

The experimental protocol complied with the tenets of the Declaration of Helsinki was approved by the French ethic committee (Comité de Protection des Personnes EST-I, numéro SI: 24.00651.000276) as required by French legislation. The ethics committee approved the study on 11 April 2024. This clinical trial is registered on ClinicalTrials.gov (reference: **NCT06438432**) and was developed in accordance with SPIRIT guidelines (60). Further information on enrollment, interventions and assessments in **Fig 1**.

**Fig 1.**
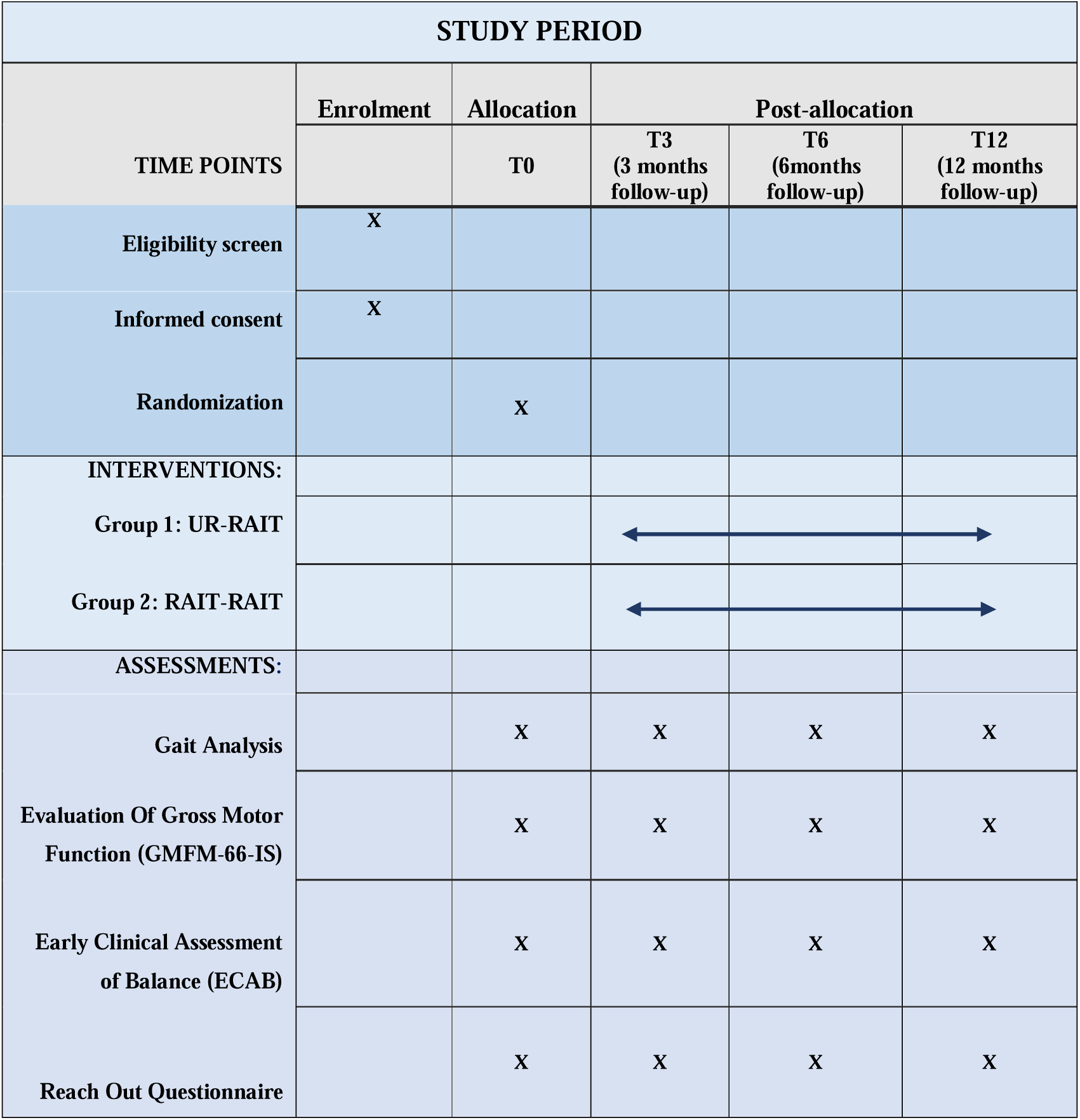
Time schedule of enrolment, interventions, and assessments. Overview of enrollment, intervention, and outcome assessments for participants, in accordance with SPIRIT 2013 guideline recommendations.

The anonymity of participants will be ensured by assigning them a unique alphanumeric reference. This reference will be used to design the participant in any files containing the data to be analyzed. Only the principal investigator and his first collaborator (CB, SZ) have access to the folder containing the conversion key. In practice, the security of the anonymity of participants is not possible due to the close collaboration between the health care professionals and the patients. However, quantitative results will be presented in accumulated form, and care will be taken to guarantee that no participants are recognizable in the results. All data from assessments and rehabilitation participation will be securely stored and the patients deleted after completion of the study.

### 2.2 Participant enrolment and study timeline

The children are recruited and evaluated at the Centre de Médecine Physique et de Réadaptation de l’Enfance (CMPRE) in Flavigny-sur-Moselle, Lorraine, France. This is a regional center providing a range of functional assessment and treatment services for children with orthopedic, neurological or neuro-orthopedic abnormalities. The children with CP recruited may be part of cohorts of patients followed at the CMPRE or be referred by pediatricians, neuro-pediatricians and physical medicine and rehabilitation physicians from different networks interacting with the CMPRE. During an initial medical consultation with a physician working in collaboration with the CMPRE, if a child is eligible, their parents are informed about this study, receive an information and consent form, and are referred to the CMPRE for an assessment of their motor skills and a medical consultation. The physiotherapist treating their child will then be informed about the study and invited to collaborate. During the second medical consultation with the principal investigator (CB) at the CMPRE, the child will be offered the opportunity to participate in the study, if their eligibility is confirmed and if the physiotherapist and parents have given their written consent. If the child is enrolled, the therapeutic group assignment contained in a sealed envelope is revealed by the principal investigator (CB) and the motor skills assessments from that day are used as baseline assessments (M0).

For children with TD, recruitment will be conducted through an email announcement presenting the study and including the information and consent form. The email will be sent to staff at the Regional Rehabilitation Institute, the Nancy Regional and University Hospital Center, and the University of Lorraine, who reside in the Nancy metropolitan area and have no hierarchical relationship with the study investigators. As with the children with CP, they will be seen in consultation by an investigating physician.

The duration of patient participation is 12 months. Patient enrolment started on 25 April 2024 and will end on 31 October 2026. This enrolment period of 30 months is in line with the usual recruitment and follow-up of children with CP by the CMPRE. The total planned duration of the study (participation and data analysis) is 54 months.

### 2.3 Study design

This protocol is a monocentric open-label randomized cross over study with two groups of children with CP pre-per and post interventional rehabilitation. The control group of children with TD will be evaluated once and compared to children with PC at their initial period of evaluation (M0), carried out after recruitment. Children with CP are followed for 12 months, with evaluations at 3, 6 and 12 months after M0.

Children are randomly divided into two groups: In the first group (UR-RAIT), UR received by children is continued during the first 3 months, and is replaced by RAIT for the following 9 months. In the second group (RAIT-RAIT), children benefit from RAIT for the entire duration of the study. An individual random allocation of participants into the two groups in blocks of four using Matlab software was carried out in advance, then placed in sealed, numbered envelopes to be used in the order of inclusion. The study design is depicted in **Fig 2**.

**Fig 2.**
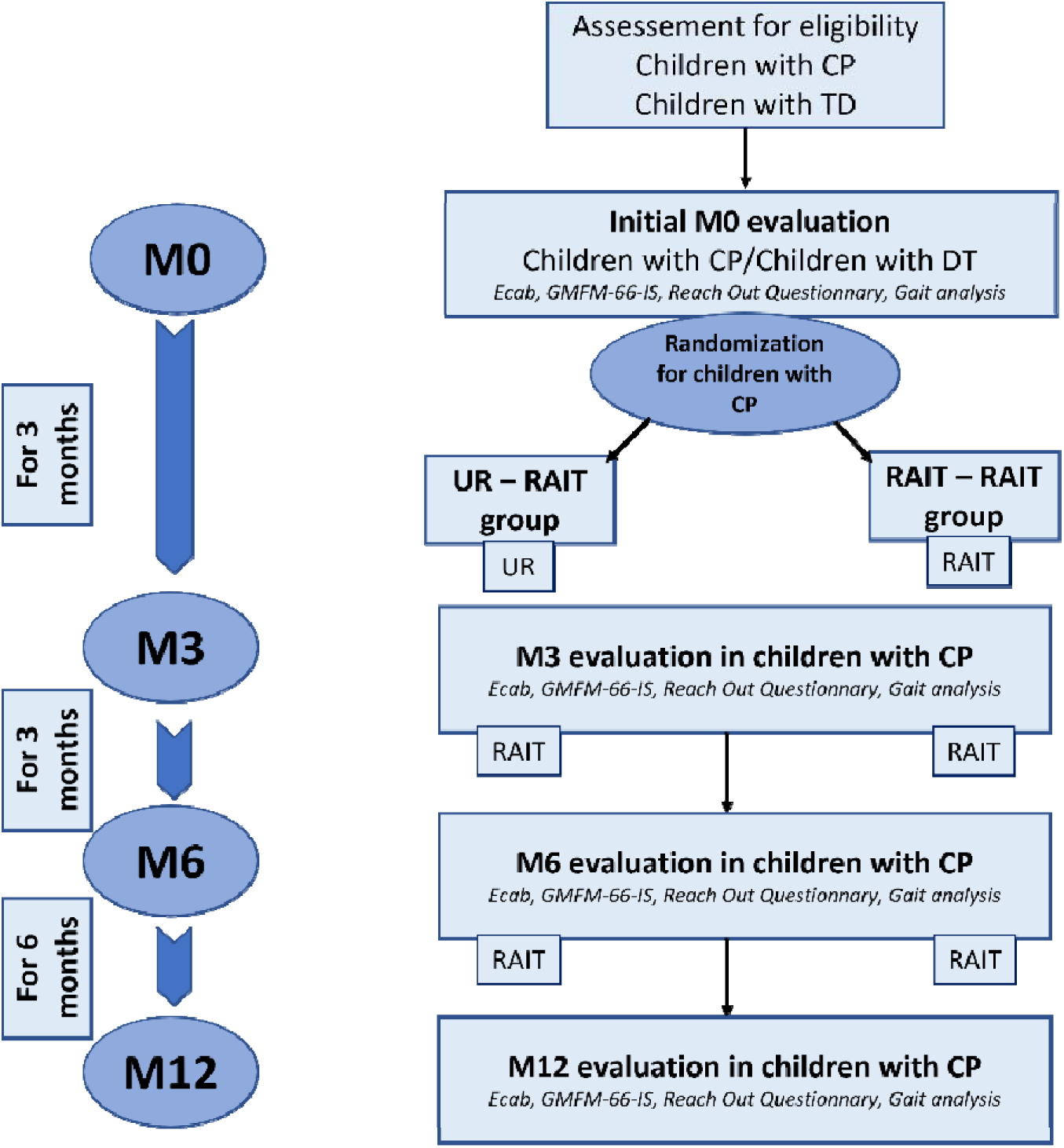
Study design. Children with cerebral palsy (CP) are randomly allocated UR – RAIT or RAIT – RAIT group. In the UR – RAIT group, Usual Rehabilitation (UR) is applied for 3 months and the Rehabilitation by activities involving the Trunk (RAIT) for the following 9 months. In the RAIT – RAIT group, RAIT is applied for 12 months. The children with CP are evaluated before the start of the rehabilitation program (at M0), after 3 months (M3), 6 months (M6) and 12 months (M12) of rehabilitation. A group of children with TD is evaluated once for comparison with the children with CP at M0.

### 2.4 Participants

#### 2.4.1 Children with CP

The main inclusion criteria for the children with CP are as follows: age between 18 months and 5 years and six months (toddlers and early childhood); the ability to walk without aids on 7 meters, i.e. the length of the walkway equipped with pressure sensors (Gross Motor Function Classification System level: I or II) (61); little or no contracture of the triceps surae (defined as forced ankle dorsiflexion of at least 5° with the knee extended), and the presence of soleus spasticity, according to the Tardieu scale (62).

The main exclusion criteria are as follows: botulinum toxin injections in the lower limbs in the 6 months preceding the study; lower limb surgery in the 12 months preceding the study; any changes in physical or orthopedic therapy within the previous 2 months; minimum hip flexion greater than 20°; pain in the lower limbs when standing or walking; insufficient cognitive level and cooperation to perform the evaluations or rehabilitation (mimic the tasks, walk as usual).

#### 2.4.2 Children with TD

The children with TD will be age-matched with the children with TD. They must be able to walk independently at 18 months of age, to have sufficient cognitive level and cooperation to perform the evaluations, with no history of neurological or orthopedic disease, no history of lower limb surgery, and no pain.

### 2.5 Evaluations

#### 2.5.1 Gait analysis

Gait analysis is threefold: analysis of trunk accelerations using IMUs, analysis of temporospatial gait parameters and centers of pressure using a walkway equipped with pressure sensors, and analysis of the Edinburgh visual gait score using video recording.

##### 2.5.1.1 Trunk dynamics

Trunk accelerations are measured by Inertial Measurement Units (IMUs), from the mTest3 system (mHealth Technologies, Bologna, Italy). They are attached to the skin by a hypoallergenic double-sided adhesive on the sternum (upper trunk), on the waist at the level of the L5 vertebra (lower trunk location close to the body center of mass) and on feet to determine foot contact and foot-off. The IMUs have a sampling frequency of 100Hz for the accelerometer signal. Data are processed using a custom-made script on Matlab R2022b (MathWorks, Inc., Natick, MA, USA). The raw data are filtered using a 10 Hz low-pass filter and averaged over all gait cycles.

Peak anterior deceleration of the sternum during WA is the primary outcome of the study. High value of this variable is suggested to reflect the higher need to break the upper trunk forward progression during WA in order to compensate for insufficient postural control of the trunk (52). Peak downward deceleration of L5 during WA is a secondary outcome: high value is suggested to reflect the higher need to break the downward movement of body center of mass during WA in order to compensate for insufficient balance and trunk control (52,63).

##### 2.5.1.2 Temporospatial parameters and trajectory of CoP

Gait pattern related to balance disorder and to toe walking are obtained by a Zeno Walkway Gait Analysis System ®) equipped with pressure sensors positioned on its surface (427 cm x 122 cm, sampling frequency of 120 Hz). These sensors detect plantar pressures during walking, allowing to assess foot prints, trajectory of CoP and temporospatial parameters, processed by PKMAS software (ProtoKinetics Movement Analysis Software). The children are asked to walk barefoot back and forth along the 7 meters-long track several times, at spontaneous speed. Series of successive gait cycles during which the child is not distracted and walks in line with the treadmill are selected in order to obtain a total of more than 17 gait cycles for each side.

The enhanced gait variability index (eGVI) is hypothesized to be increased. It is a composite score based on 9 temporospatial parameters that quantifies the distance between the amount of variability observed in an asymptomatic reference group and the amount of variability observed in the patient (64,65). Indeed, this index that assesses instability during gait and the risk of falling is usually high in children with CP, as they are impacted by balance disorders (66,67).

Step width is hypothesized to increase. Indeed, this variable reflects a strategy for reducing the risk of falls in unstable gait and is usually high in children with CP (68).

Center of pressure location normalized to foot length during WA, on the affected side in cases of hemiplegic CP or on both sides in cases of paraplegic CP, is hypothesized to be excessively shifted toward the front of the foot due to toe walking, with a higher anterior location in cases of more important toe walking (69).

##### 2.5.1.3 Visual evaluation of the gait kinematic pattern

The Edinburgh Visual Gait Score, a kinematic gait pattern score based on two cameras recording front and side view, is hypothesized to be decreased as the gait pattern in CP children, including toe walking, is different from children with TD (56).

#### 2.5.2 Evaluation of gross motor function by clinical scores

The Item Set of the Gross Motor Function Measurement 66 (GMFM-66-IS) is hypothesized to be reduced with CP. Compared to the standardized 66-items used to assess gross motor function (57), the item set version (GMFM-66-IS) which has the same validity is faster to realize (around 20 to 30 minutes versus 60 to 80 minutes) since it uses 15 to 39 items selected according to the achievement of 3 main items (70). Thus, the GMFM-66-IS by saving time is of great interest for the young children of this study.

The Early Clinical Assessment of Balance (ECAB) is hypothesized to be reduced in children with CP. This 13-item clinical scale assesses postural stability (balance ability) in children with cerebral palsy, with two subscales: one dedicated to head and trunk postural control, the other to sitting and standing postural control. This scale is validated for children aged 1.5 to 11 years, regardless of GMFCS level (58,71). The optimal score is 100.

#### 2.5.3 Evaluation of upper limb function, activity and participation

Evaluation of upper limb function and child’s activity and participation is made by the Reach Out Questionnaire. This questionnaire filled by the parents ensure a holistic overview of functioning, when a hand and upper limb are affected, including the assessment of activity limitations, participation in activities, attitudes in different environments and satisfaction, according to the domains of the ICF (59). Rehabilitation by activities involving the trunk is hypothesized to improve hand and upper limb function in case of disorders affecting the upper limb function and the child’s participation, since the trunk appears central in motor activities.

Therefore, by combining neuromuscular assessment, evaluation of overall body and trunk postural control, overall motor function, walking, as well as reaching, grasping behavior, and participation, our study evaluates the effect of RAIT on all ICF domains.

### 2.6 Intervention

If the children are randomly assigned to the first group (UR-RAIT), they will receive 3 months of UR followed by RAIT for the following 9 months. In the second group (RAIT-RAIT), children benefit from RAIT for the entire duration of the study.

#### 2.6.1 Rehabilitation by activities involving the trunk

If the physiotherapist caring for the child with CP agrees to participate in the study, he/she will be initially briefed on the principles of RAIT and given detailed instructions on its content shortly before the start of the RAIT treatment.

Rehabilitation by activities involving the trunk (RAIT) exploits the automatic postural control of body balance, an essential aspect of motor function that enables individuals to maintain balance during static postures or motor activities. It is also noteworthy that our rehabilitation protocol extends over a duration of up to one year, which appears to be an important factor for maintaining improvements in motor function. In our previous study in children with CP aged from 5 to 12 years, the RAIT was applied for 3 months with significant improvement of gait dynamics and pattern (52).

The RAIT program focuses on self-directed exercises actively performed by the child, so that balance is controlled by the child and not by another person, in order to develop and engram balance strategies. These exercises aim to enhance body postural control and balance, including the trunk and affected muscles, rather than focusing rehabilitation on the affected muscles. In fact, RAIT does not only involve the trunk, but the entire body in selected intermediate postures involving the trunk that the child performs on his/her own. These postural activities are less challenging than standing or walking when considering balance control and help eliminate fear of falling. These activities allow automatic recruitment of deficient muscles through their contribution to balance control. For example, children with hemiplegic CP often have limited voluntary control of their affected upper limb: using the latter as support will automatically activate wrist and elbow extension and shoulder control without voluntary control of the affected muscles.

Self-exercises are performed by the child, under the guidance of the physiotherapist, in separate small sessions, if possible, for a total of 20 to 30 minutes a day, 6 days a week, at home under the supervision of a parent and by the physiotherapist in one to two sessions per week. The use of a monitoring log, in which the physiotherapist notes down the exercises to be performed and the parents indicate those that have been completed, will allow for smoother and more regular monitoring of difficulties and progress in rehabilitation **(Fig 3).**

**Fig 3.**
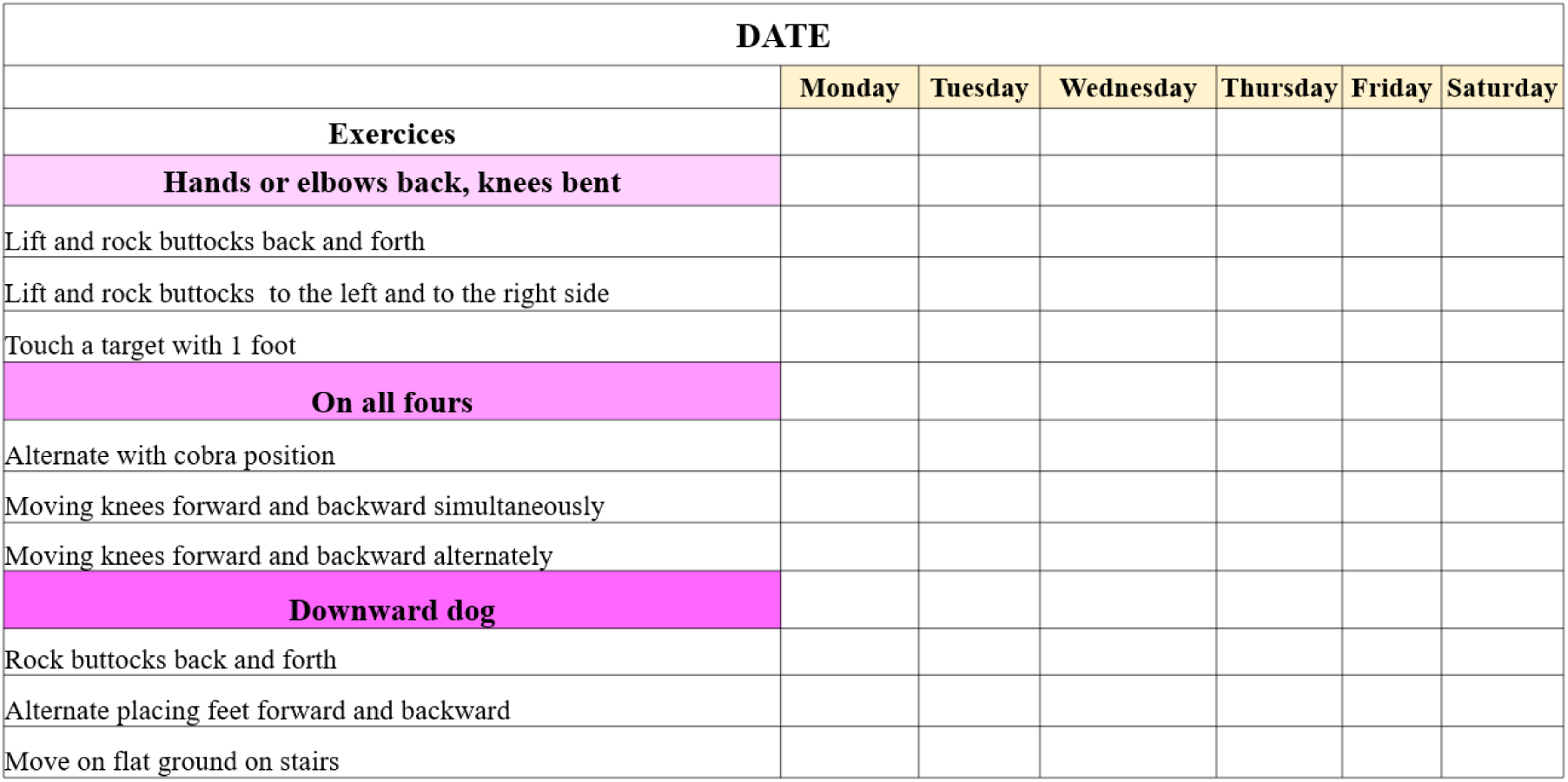
Part of a monitoring log in which a series of exercises are suggested. The physical therapist selects and adjusts some of them according to the child’s abilities, and the parents indicate which ones have been completed by the child.

#### 2.6.2 Usual rehabilitation

The UR corresponds to the type of rehabilitation already received by the child before the study. It variously combines muscle stretching, muscle strengthening (e.g., resistance training), muscle tone reduction (e.g., Bobath concept neuro – developmental treatment), and upper and lower limb motor skill training facilitated by the therapist. These therapies usually involve limited groups of muscles in elementary stretching or actions. In the UR-RAIT group, the physiotherapist is asked to give to the parents an adapted selection of the usual rehabilitation to be achieved at home for a total duration of 20 to 30 minutes per day, the days without physiotherapist session, for a total of 6 days a week. A monitoring log, as previously described for RAIT, will be used with the exercises prescribed by the physiotherapist.

### 2.7 Data management

The study is conducted in accordance with the provisions of Law No. 78-17 of January 6, 1978, on information technology, files, and civil liberties, as amended by Law No. 2018-493 of June 20, 2018, on the protection of personal data, and European Regulation No. 2016/679 (GDPR). The reference methodology for the processing of personal data in the context of biomedical research applied in this study is MR-001.

Medical data concerning the children, and data necessary for the study will be collected and transmitted under the responsibility of the Research Promotor and its Data Protection Officer. The data collected during this study may be used for publication in scientific journals, and may be reused for other research in the field of trunk control and gait disorders in CP.

#### 2.7.1 Sample size

The sample size is calculated for the main criterion. The distribution of peak anterior deceleration of the sternum during WA of gait in a previous study followed a normal distribution (52). On average, this distribution was centered on 3.0 ± 1.1 m. s-2 (mean ± 1 standard deviation) in children with CP and on 1.1 ± 0.3 m. s-2 in children with DT. The expected effect of RAIT is a minimal reduction in peak anterior deceleration of the sternum during WA by one-third of its value, similar to the effect of RAIT in the previous study (52). In order to show an effect of RAIT using a repeated-measures ANOVA in which we expect a medium effect size, with an alpha threshold set at 0.05 and a power of 80%, a total number of 24 patients, i.e., 12 per group, is required (calculation using G*Power 3.1 software). In addition, to account for the greater variability of gait dynamics in toddlers and early childhood compared to middle childhood (72) and possible lost to follow-up, we plan to recruit 32 PC children (16 per group). Missing data for the primary and secondary outcomes will be handled under a missing-at-random assumption using multiple imputation by chained equations. Input datasets will be created including baseline outcome, treatment group, participation in rehabilitation activities, age and sex as predictors.

### 2.8 Statistical analysis

Statistical analyses will be carried out within the framework of general linear models, the overall sample (N=64) and for each group (n=32) being a priori sufficient for the application of such procedures. The conditions for applying such tests will be checked beforehand, in particular the normal distribution (using quantile-quantile graphs and Shapiro-Wilk tests) and the homogeneity of variance (Levene tests). If the data do not meet these conditions, non-parametric equivalents will be used.

To compare the children with CP vs. the children with TD, independent samples t-tests will be used for each outcome variable, checked of normality and the homogeneity of variance. In case of normality violation, the non-parametric Mann–Whitney U test will be applied. In children with CP, taken together at M0, correlation tests will be performed using Pearson’s coefficient to test correlations between variables related to trunk dynamics, gait instability, and gross motor control.

To compare the groups UR-RAIT and RAIT-RAIT, a repeated measures ANOVA will be conducted. The between-subject factor is the rehabilitation group (UR-RAIT vs. RAIT-RAIT), and the within-subject factor is time, assessed across four periods: 0 months (M0), 3 months (M3), 6 months (M6), and 12 months (M12). If assumptions of normality or sphericity is violated, appropriate corrections (such as Greenhouse-Geisser) or non-parametric alternatives will be applied (Friedman’s test).

If the repeated measures ANOVA is significant, analysis of local comparisons using post-hoc analyses (Holm or Bonferroni) will be conducted to further explore the results. Comparisons between evaluations (M0, M3, M6, and M12) will be performed to examine the evolution of outcomes over time within each group. In the case of a significant group × time interaction, simple effects analyses will be carried out to compare the two groups (UR-RAIT vs. RAIT-RAIT) at each time point, as well as to assess the progression within each group across the different time intervals.

This analysis will allow us to explore three key aspects: (1) the compared effectiveness of both rehabilitation approaches, between M0 and M3 for the primary outcome (peak anterior deceleration of the sternum during WA) and the secondary outcomes; (2) the reproducibility of the RAIT effect, by observing whether similar improvements occur between M3 and M6 in the UR-RAIT group and between M0 and M3 in the RAIT-RAIT group; and (3) the amplification of the RAIT effect with continued application, by comparing changes between M6 and M0 versus M3 and M0, and between M12 and M0 versus M6 and M0 in the RAIT-RAIT group, as well as between M12 and M3 versus M6 and M3 in the UR-RAIT group.

## 3. Discussion

Due to the high frequency of balance and postural disorders in children with CP (73) and in particular of trunk disorders (29,30,42), which affect the different domains of the ICF-CY (6,74,75), it is important to offer adequate rehabilitation programs targeting these specific deficits. Among trunk targeted interventions that develop for more than a decade (76–78), to our knowledge, this study self-distinguish by the original approach and content of RAIT, by the young age of the children with CP and by the long-term application and evaluation of RAIT.

The RAIT considers balance control as a priority motor function that significantly impacts motor patterns and activities in upper motor neuron syndromes such as in cerebral palsy (79). In these pathologies, the brainstem and cerebellum that support balance and postural control are usually anatomically intact (79). In RAIT, the children perform functional exercises, i.e., self-directed activities based on intermediate postures involving the trunk and the four limbs and exploiting the automatic control of balance. In other words, the affected trunk and limbs will automatically participate to balance control according to their involvement to balance in intermediate postures. The intermediate postures are chosen to be securely performed with no fear of falling and to further involve the affected muscles in body balance. For example, being on all fours or in cobra posture would automatically recruit extensors of the different joints of the upper limbs to contribute to body balance **(see Fig. 2)**. Activities in progressively more challenging intermediate postures for balance will be chosen to further increase the contribution of the trunk and affected limbs to balance in various motor activities. Therefore, an improvement in activities and participation according to the ICF-CY is expected after RAIT. The self-exercises of RAIT, carried out daily, are subject to individualized adjustments in line with the child’s abilities.

In addition, this study could provide new elements towards a rethinking of the pathophysiology of motor disorders during walking in children with CP. For example, recent studies question spasticity as the main explanatory factor for toe walking in children with CP (47,80). One possible explanation is that it is an adaptive strategy (46,48) to compensate for a deficit in trunk control during walking. Indeed, the improvement in trunk control through RAIT is expected to lead to a reduction in compensation exerted by the foot: a reduction in the anterior deceleration of the sternum is expected to be associated to a reduction in the anterior shift of the CoP, during the WA of walking.

### 3.1 Limitations

This study presents several limitations mainly related to the young population of children with CP.

#### 3.1.1 Participation

In longitudinal studies, involving young children can be particularly challenging, as fluctuations in the child’s level of cooperation, alertness or health may prevent regular involvement or completion of the rehabilitation program 6 days per week. Additionally, participation rates may be reduced by time constraints due to parents’ or school schedules.

#### 3.1.2 Developmental variability

At young age (18 months to 5 and a half years of age), children’s development is highly variable, either for motor, cognitive, emotional, or social development. This natural variability linked to the young age of the children, combined with the variability in the severity of CP and the performance of RAIT, makes it difficult to estimate the number of subjects required for statistical purposes.

#### 3.1.3 Ethical considerations

When conducting research in children, informed consent must be obtained from the parents or legal guardians, and if possible, from the child. In practice, due to the young age of children, communication with the child is mainly dedicated to the good realization of the rehabilitation content.

### 3.2 Perspectives

In case of good results, RAIT, which requires regular monitoring but little equipment and resources, may be to consider for routine clinical practice in children with CP. The self-exercises proposed, guided by the physiotherapist, would be carried out at home under the supervision of parents. Indeed, the active involvement of parents in their child’s care process appears to be a considerable asset for the success of this intensive approach for motor improvement including gait quality. Given the central role of the trunk in the development of body balance control, applying RAIT in young children with CP before they experience standing and walking would constitute the next step to be studied to reduce or avoid the development of compensatory mechanism by the lower limbs during standing and walking acquisition.

## Supporting information

S1 Appendix. SPIRIT guidelines checklist. (PDF)

## Supporting information

RAIT in CP CPP protocol ENGLISH

RAIT in CP CPP ENGLISH

## Data Availability

No datasets were generated or analysed during the current study. All relevant data from this study will be made available upon study completion.

## References

1. Oskoui M, Coutinho F, Dykeman J, Jetté N, Pringsheim T. An update on the prevalence of cerebral palsy: a systematic review and metaLanalysis. Dev Med Child Neurol [Internet]. 2013 June [cited 2025 Sept 12];55(6):509–19. Available from: https://onlinelibrary.wiley.com/doi/10.1111/dmcn.12080

2. Bax M, Goldstein M, Rosenbaum P, Leviton A, Paneth N, Dan B, et al. Proposed definition and classification of cerebral palsy, April 2005. Dev Med Child Neurol [Internet]. 2005 July 14 [cited 2024 Aug 20];47(8):571–6. Available from: http://www.journals.cambridge.org/abstract_S001216220500112X

3. Rosenbaum P, Paneth N, Leviton A, Goldstein M, Bax M, Damiano D, et al. A report: the definition and classification of cerebral palsy April 2006. Dev Med Child Neurol [Internet]. 2006 [cited 2018 Nov 2];49:8–14. Available from: http://doi.wiley.com/10.1111/j.1469-8749.2007.tb12610.x

4. World Health Organization. International classification of functioning, disability and health: children and youth version: ICF-CY. 2007;322.

5. Woollacott M, Burtner P. Neural and musculoskeletal contributions to the development of stance balance control in typical children and in children with cerebral palsy. Acta Paediatr [Internet]. 1996 Oct [cited 2025 Sept 18];85(s416):58–62. Available from: https://onlinelibrary.wiley.com/doi/10.1111/j.1651-2227.1996.tb14279.x

6. Kara OK, Gursen C, Cetin SY, Tascioglu EN, Muftuoglu S, Damiano DL. The effects of power exercises on body structure and function, activity and participation in children with cerebral palsy: an ICF-based systematic review. Disabil Rehabil. 2023 Oct 23;45(22):3705–18.

7. Massion J. Postural control system. Curr Opin Neurobiol [Internet]. 1994 Dec [cited 2024 Jan 23];4(6):877–87. Available from: https://linkinghub.elsevier.com/retrieve/pii/0959438894901376

8. Massion J, Alexandrov A, Frolov A. Why and how are posture and movement coordinated? Prog Brain Res. 2004;143:13–27.

9. Assaiante C, Mallau S, Viel S, Jover M, Schmitz C. Development of postural control in healthy children: a functional approach. Neural Plast. 2005;12(2–3):109–18.

10. Velghe S, Rameckers E, Meyns P, Johnson C, Hallemans A, Verbecque E, et al. Effects of a highly intensive balance therapy camp in children with developmental coordination disorder – An intervention protocol. Res Dev Disabil [Internet]. 2024 Apr [cited 2024 Mar 29];147:104694. Available from: https://linkinghub.elsevier.com/retrieve/pii/S089142222400026X

11. Pin TW, Butler PB, Cheung HM, Shum SLF. Relationship between segmental trunk control and gross motor development in typically developing infants aged from 4 to 12 months: a pilot study. BMC Pediatr [Internet]. 2019 Dec [cited 2024 July 17];19(1):425. Available from: https://bmcpediatr.biomedcentral.com/articles/10.1186/s12887-019-1791-1

12. Butler PB. A preliminary report on the effectiveness of trunk targeting in achieving independent sitting balance in children with cerebral palsy. Clin Rehabil. 1998;12(4):281– 93.

13. de Graaf-Peters, Blauwhospers C, Dirks T, Bakker H, Bos A, Haddersalgra M. Development of postural control in typically developing children and children with cerebral palsy: Possibilities for intervention? Neurosci Biobehav Rev [Internet]. 2007 [cited 2015 Nov 24];31(8):1191–200. Available from: http://linkinghub.elsevier.com/retrieve/pii/S0149763407000486

14. Kyvelidou A, Harbourne RT, Willett SL, Stergiou N. Sitting Postural Control in Infants With Typical Development, Motor Delay, or Cerebral Palsy: Pediatr Phys Ther [Internet]. 2013 [cited 2019 Feb 20];25(1):46–51. Available from: http://content.wkhealth.com/linkback/openurl?sid=WKPTLP:landingpage&an=00001577-201325010-00013

15. Cignetti F, Chabeauti PY, Sveistrup H, Vaugoyeau M, Assaiante C. Updating process of internal models of action as assessed from motor and postural strategies in children. Neuroscience. 2013 Mar;233:127–38.

16. Rival C, Ceyte H, Olivier I. Developmental changes of static standing balance in children. Neurosci Lett. 2005 Mar;376(2):133–6.

17. Pierret J, Beyaert C, Paysant J, Caudron S. How do children aged 6 to 11 stabilize themselves on an unstable sitting device? The progressive development of axial segment control. Hum Mov Sci [Internet]. 2020 June [cited 2024 Aug 16];71:102624. Available from: https://linkinghub.elsevier.com/retrieve/pii/S0167945719303781

18. Adolph KE, Vereijken B, Shrout PE. What Changes in Infant Walking and Why. Child Dev [Internet]. 2003 Mar [cited 2024 June 18];74(2):475–97. Available from: https://srcd.onlinelibrary.wiley.com/doi/10.1111/1467-8624.7402011

19. Malina RM. Motor Development during Infancy and Early Childhood: Overview and Suggested Directions for Research. Int J Sport Health Sci [Internet]. 2004 [cited 2024 June 18];2:50–66. Available from: http://www.jstage.jst.go.jp/article/ijshs/2/0/2_0_50/_article

20. Assaiante C. Development of locomotor balance control in healthy children. Neurosci Biobehav Rev. 1998;22(4):527–32.

21. Brenière Y, Bril B. Development of postural control of gravity forces in children during the first 5 years of walking. Exp Brain Res. 1998 Aug 10;121(3):255–62.

22. Bril B, Dupuy L, Dietrich G, Corbetta D. Learning to tune the antero-posterior propulsive forces during walking: a necessary skill for mastering upright locomotion in toddlers. Exp Brain Res. 2015 Oct;233(10):2903–12.

23. Cappozzo A. The forces and couples in the human trunk during level walking. J Biomech [Internet]. 1983 Jan [cited 2023 Dec 12];16(4):265–77. Available from: https://linkinghub.elsevier.com/retrieve/pii/0021929083901343

24. Cromwell RL, Aadland-Monahan TK, Nelson AT, Stern-Sylvestre SM, Seder B. Sagittal Plane Analysis of Head, Neck, and Trunk Kinematics and Electromyographic Activity During Locomotion. J Orthop Sports Phys Ther. 2001 May;31(5):255–62.

25. Honeine JL, Schieppati M, Gagey O, Do MC. By counteracting gravity, triceps surae sets both kinematics and kinetics of gait. Physiol Rep. 2014 Feb;2(2):e00229.

26. Kavanagh J, Barrett R, Morrison S. The role of the neck and trunk in facilitating head stability during walking. Exp Brain Res. 2006 July;172(4):454–63.

27. Winter DA. Human balance and posture control during standing and walking. Gait Posture. 1995;3(4):193–214.

28. Brogren E, Hadders-Algra M, Forssberg H. Postural Control in Sitting Children with Cerebral Palsy. Neurosci Biobehav Rev [Internet]. 1998 Mar [cited 2024 Jan 19];22(4):591–6. Available from: https://linkinghub.elsevier.com/retrieve/pii/S0149763497000493

29. Heyrman L, Desloovere K, Molenaers G, Verheyden G, Klingels K, Monbaliu E, et al. Clinical characteristics of impaired trunk control in children with spastic cerebral palsy. Res Dev Disabil [Internet]. 2013 Jan [cited 2016 May 12];34(1):327–34. Available from: http://linkinghub.elsevier.com/retrieve/pii/S0891422212002211

30. Pierret J, Caudron S, Paysant J, Beyaert C. Impaired postural control of axial segments in children with cerebral palsy. Gait Posture [Internet]. 2021 Mar 8 [cited 2021 Mar 9]; Available from: https://www.sciencedirect.com/science/article/pii/S0966636221000977

31. Saavedra SL, Woollacott MH. Segmental Contributions to Trunk Control in Children With Moderate-to-Severe Cerebral Palsy. Arch Phys Med Rehabil [Internet]. 2015 June [cited 2024 Jan 17];96(6):1088–97. Available from: https://linkinghub.elsevier.com/retrieve/pii/S0003999315000921

32. Attias M, Bonnefoy-Mazure A, Lempereur M, Lascombes P, De Coulon G, Armand S. Trunk movements during gait in cerebral palsy. Clin Biomech [Internet]. 2015 Jan [cited 2024 Jan 17];30(1):28–32. Available from: https://linkinghub.elsevier.com/retrieve/pii/S0268003314002903

33. Hazari A, Agouris I, Wakode PS, Jadhav RA, Sharma N, Jena S, et al. Head and trunk kinematics and kinetics in normal and cerebral palsy gait: a systematic review. Eur J Physiother [Internet]. 2020 May 3 [cited 2024 June 18];22(3):168–77. Available from: https://www.tandfonline.com/doi/full/10.1080/21679169.2019.1573919

34. Kiernan D. The relationship of trunk kinematics and kinetics with lower limb pathology during gait in children with spastic cerebral palsy. Gait Posture [Internet]. 2021 May [cited 2024 July 10];86:33–7. Available from: https://linkinghub.elsevier.com/retrieve/pii/S0966636221000825

35. Leardini A, Biagi F, Merlo A, Belvedere C, Benedetti MG. Multi-segment trunk kinematics during locomotion and elementary exercises. Clin Biomech [Internet]. 2011 July [cited 2024 Jan 24];26(6):562–71. Available from: https://linkinghub.elsevier.com/retrieve/pii/S0268003311000386

36. Wallard L, Dietrich G, Kerlirzin Y, Bredin J. Balance control in gait children with cerebral palsy. Gait Posture [Internet]. 2014 May [cited 2018 June 6];40(1):43–7. Available from: http://linkinghub.elsevier.com/retrieve/pii/S0966636214000733

37. Katz-Leurer M, Rotem H, Keren O, Meyer S. Balance abilities and gait characteristics in post-traumatic brain injury, cerebral palsy and typically developed children. Dev Neurorehabilitation [Internet]. 2009 Jan [cited 2019 Aug 28];12(2):100–5. Available from: http://www.tandfonline.com/doi/full/10.1080/17518420902800928

38. Kim CJ, Son SM. Comparison of Spatiotemporal Gait Parameters between Children with Normal Development and Children with Diplegic Cerebral Palsy. J Phys Ther Sci [Internet]. 2014 [cited 2020 Jan 12];26(9):1317–9. Available from: http://jlc.jst.go.jp/DN/JST.JSTAGE/jpts/26.1317?lang=en&from=CrossRef&type=abstrac t

39. Saether R, Helbostad JL, Adde L, Brændvik S, Lydersen S, Vik T. Gait characteristics in children and adolescents with cerebral palsy assessed with a trunk-worn accelerometer. Res Dev Disabil [Internet]. 2014 July [cited 2024 Feb 28];35(7):1773–81. Available from: https://linkinghub.elsevier.com/retrieve/pii/S0891422214000791

40. Summa A, Vannozzi G, Bergamini E, Iosa M, Morelli D, Cappozzo A. Multilevel Upper Body Movement Control during Gait in Children with Cerebral Palsy. Lebedev MA, editor. PLOS ONE [Internet]. 2016 Mar 21 [cited 2025 July 8];11(3):e0151792. Available from: https://dx.plos.org/10.1371/journal.pone.0151792

41. Heyrman L, Feys H, Molenaers G, Jaspers E, Monari D, Nieuwenhuys A, et al. Altered trunk movements during gait in children with spastic diplegia: Compensatory or underlying trunk control deficit? Res Dev Disabil [Internet]. 2014 Sept [cited 2023 Nov 10];35(9):2044–52. Available from: https://linkinghub.elsevier.com/retrieve/pii/S0891422214001991

42. Saether R, Helbostad JL, Adde L, Braendvik S, Lydersen S, Vik T. The relationship between trunk control in sitting and during gait in children and adolescents with cerebral palsy. Dev Med Child Neurol [Internet]. 2015 Apr [cited 2017 Nov 2];57(4):344–50. Available from: http://doi.wiley.com/10.1111/dmcn.12628

43. Balzer J, Marsico P, Mitteregger E, van der Linden ML, Mercer TH, van Hedel HJA. Influence of trunk control and lower extremity impairments on gait capacity in children with cerebral palsy. Disabil Rehabil. 2017 Sept 24;1–7.

44. Armand S, Watelain E, Mercier M, Lensel G, Lepoutre FX. Identification and classification of toe-walkers based on ankle kinematics, using a data-mining method. Gait Posture [Internet]. 2006 Feb [cited 2024 July 9];23(2):240–8. Available from: https://linkinghub.elsevier.com/retrieve/pii/S0966636205000317

45. Worthen-Chaudhari L, Bing J, Schmiedeler JP, Basso DM. A new look at an old problem: Defining weight acceptance in human walking. Gait Posture [Internet]. 2014 Jan [cited 2025 July 19];39(1):588–92. Available from: https://linkinghub.elsevier.com/retrieve/pii/S0966636213006206

46. Beyaert C, Pierret J, Vasa R, Paysant J, Caudron S. Toe walking in children with cerebral palsy: a possible functional role for the plantar flexors. J Neurophysiol [Internet]. 2020 Oct 1 [cited 2024 Jan 25];124(4):1257–69. Available from: https://journals.physiology.org/doi/10.1152/jn.00717.2019

47. Willerslev-Olsen M, Andersen JB, Sinkjaer T, Nielsen JB. Sensory feedback to ankle plantar flexors is not exaggerated during gait in spastic hemiplegic children with cerebral palsy. J Neurophysiol. 2014 Feb;111(4):746–54.

48. Lorentzen J, Willerslev-Olsen M, Hüche Larsen H, Farmer SF, Nielsen JB. Maturation of feedforward toe walking motor program is impaired in children with cerebral palsy. Brain [Internet]. 2019 Mar 1 [cited 2019 May 29];142(3):526–41. Available from: https://academic.oup.com/brain/article/142/3/526/5306635

49. Correa TA, Schache AG, Graham HK, Baker R, Thomason P, Pandy MG. Potential of lower-limb muscles to accelerate the body during cerebral palsy gait. Gait Posture [Internet]. 2012 June [cited 2024 Feb 27];36(2):194–200. Available from: https://linkinghub.elsevier.com/retrieve/pii/S0966636212000653

50. Honeine JL, Schieppati M, Gagey O, Do MC. The Functional Role of the Triceps Surae Muscle during Human Locomotion. Kreplak L, editor. PLoS ONE [Internet]. 2013 Jan 16 [cited 2024 Jan 5];8(1):e52943. Available from: https://dx.plos.org/10.1371/journal.pone.0052943

51. Neptune RR, Kautz SA, Zajac FE. Contributions of the individual ankle plantar flexors to support, forward progression and swing initiation during walking. J Biomech [Internet]. 2001 Nov [cited 2024 Jan 31];34(11):1387–98. Available from: https://linkinghub.elsevier.com/retrieve/pii/S0021929001001051

52. Pierret, J., Beyaert, C., Vasa, R., Rumilly, E., Paysant, J., & Caudron, S. (2023). Rehabilitation of Postural Control and Gait in Children with Cerebral Palsy: The Beneficial Effects of Trunk-Focused Postural Activities. Developmental Neurorehabilitation, 26(3), 180–192. 10.1080/17518423.2023.2193269

53. Santamaria V, Ai X, Chin K, Dutkowsky JP, Gordon AM, Agrawal SK. Study protocol for a randomised controlled trial to determine the efficacy of an intensive seated postural intervention delivered with robotic and rigid trunk support systems. BMJ Open [Internet]. 2023 Aug [cited 2024 June 21];13(8):e073166. Available from: https://bmjopen.bmj.com/lookup/doi/10.1136/bmjopen-2023-073166

54. Munaf A, Mehboob S, Razzaq M, Younas M, Umair S, Waseem I, et al. Effect of Trunk Exercises on Trunk Control, Balance, and Mobility Function in Children with Hemiparetic CP. Pak J Med Health Sci [Internet]. 2022 Nov 30 [cited 2024 Feb 5];16(11):95–8. Available from: https://pjmhsonline.com/index.php/pjmhs/article/view/3263

55. Abidin N, Ünlü Akyüz E, Cankurtaran D, Karaahmet ÖZ, Tezel N. The effect of robotic rehabilitation on posture and trunk control in non-ambulatory cerebral palsy. Assist Technol [Internet]. 2024 Nov [cited 2025 July 8];36(6):422–8. Available from: https://www.tandfonline.com/doi/full/10.1080/10400435.2022.2059592

56. Read HS, Hazlewood ME, Hillman SJ, Prescott RJ, Robb JE. Edinburgh Visual Gait Score for Use in Cerebral Palsy: J Pediatr Orthop [Internet]. 2003 May [cited 2024 Aug 19];23(3):296–301. Available from: http://journals.lww.com/01241398-200305000-00005

57. Russell DJ, Avery LM, Rosenbaum PL, Raina PS, Walter SD, Palisano RJ. Improved Scaling of the Gross Motor Function Measure for Children With Cerebral Palsy: Evidence of Reliability and Validity. Phys Ther [Internet]. 2000 Sept 1 [cited 2023 Apr 4];80(9):873–85. Available from: https://academic.oup.com/ptj/article/80/9/873/2842507

58. McCoy SW, Bartlett DJ, Yocum A, Jeffries L, Fiss AL, Chiarello L, et al. Development and validity of the early clinical assessment of balance for young children with cerebral palsy. Dev Neurorehabilitation [Internet]. 2014 Dec [cited 2024 Mar 7];17(6):375–83. Available from: http://www.tandfonline.com/doi/full/10.3109/17518423.2013.827755

59. Ma Y, Aslam R, Jester A. Development and validation of a WHO ICF compliant hand and upper limb assessment tool for children: The Reach Out questionnaire. J Hand Ther [Internet]. 2023 Oct [cited 2024 Mar 8];36(4):1000–6. Available from: https://linkinghub.elsevier.com/retrieve/pii/S0894113023000340

60. Chan AW, Tetzlaff JM, Altman DG, Laupacis A, Gøtzsche PC, Krleža-Jerić K, et al. SPIRIT 2013 Statement: Defining Standard Protocol Items for Clinical Trials. Ann Intern Med [Internet]. 2013 Feb 5 [cited 2025 Sept 15];158(3):200–7. Available from: https://www.acpjournals.org/doi/10.7326/0003-4819-158-3-201302050-00583

61. Palisano R, Rosenbaum P, Walter S, Russell D, Wood E, Galuppi B. Development and reliability of a system to classify gross motor function in children with cerebral palsy. Dev Med Child Neurol [Internet]. 2008 Sept 29 [cited 2018 Nov 2];39(4):214–23. Available from: http://doi.wiley.com/10.1111/j.1469-8749.1997.tb07414.x

62. Yelnik A, Albert T, Bonan I, Laffont I. A Clinical Guide to Assess the Role of Lower Limb Extensor Overactivity in Hemiplegic Gait Disorders. Stroke [Internet]. 1999 Mar [cited 2025 Sept 4];30(3):580–5. Available from: https://www.ahajournals.org/doi/10.1161/01.STR.30.3.580

63. Brenière Y, Bril B. Development of postural control of gravity forces in children during the first 5 years of walking. Exp Brain Res [Internet]. 1998 Aug 10 [cited 2025 Sept 1];121(3):255–62. Available from: http://link.springer.com/10.1007/s002210050458

64. Gouelle A. Use of Functional Ambulation Performance Score as measurement of gait ability: Review. J Rehabil Res Dev [Internet]. 2014 [cited 2024 Feb 15];51(5):665–74. Available from: http://www.rehab.research.va.gov/jour/2014/515/pdf/JRRD-2013-09-0198.pdf

65. Gouelle A, Rennie L, Clark DJ, Mégrot F, Balasubramanian CK. Addressing limitations of the Gait Variability Index to enhance its applicability: The enhanced GVI (EGVI). Tan MP, editor. PLOS ONE [Internet]. 2018 June 1 [cited 2024 Feb 28];13(6):e0198267. Available from: https://dx.plos.org/10.1371/journal.pone.0198267

66. Joanna M, Magdalena S, Katarzyna BM, Daniel S, Ewa LD. The Utility of Gait Deviation Index (GDI) and Gait Variability Index (GVI) in Detecting Gait Changes in Spastic Hemiplegic Cerebral Palsy Children Using Ankle–Foot Orthoses (AFO). Children [Internet]. 2020 Sept 25 [cited 2023 Dec 7];7(10):149. Available from: https://www.mdpi.com/2227-9067/7/10/149

67. Prosser LA, Aguirre MO, Zhao S, Bogen DK, Pierce SR, Nilan KA, et al. Infants at risk for physical disability may be identified by measures of postural control in supine. Pediatr Res [Internet]. 2022 Apr [cited 2025 Mar 4];91(5):1215–21. Available from: https://www.nature.com/articles/s41390-021-01617-0

68. Kurz MJ, Arpin DJ, Corr B. Differences in the dynamic gait stability of children with cerebral palsy and typically developing children. Gait Posture [Internet]. 2012 July [cited 2018 June 18];36(3):600–4. Available from: http://linkinghub.elsevier.com/retrieve/pii/S0966636212001919

69. Mancinelli C, Patel S, Deming LC, Schmid M, Patritti BL, Chu JJ, et al. Assessing the feasibility of classifying toe-walking severity in children with cerebral palsy using a sensorized shoe. In: 2009 Annual International Conference of the IEEE Engineering in Medicine and Biology Society [Internet]. Minneapolis, MN: IEEE; 2009 [cited 2023 Sept 13]. p. 5163–6. Available from: http://ieeexplore.ieee.org/document/5332733/

70. Avery LM, Russell DJ, Raina PS, Walter SD, Rosenbaum PL. Rasch analysis of the gross motor function measure: Validating the assumptions of the Rasch model to create an interval-level measure. Arch Phys Med Rehabil [Internet]. 2003 May [cited 2023 Apr 4];84(5):697–705. Available from: http://www.mosby.com/scripts/om.dll/serve?action=searchDB&searchDBfor=art&artTyp e=abs&id=as0003999303048967

71. LaForme Fiss A, McCoy SW, Bartlett D, Avery L, Hanna SE, On Track Study Team. Developmental Trajectories for the Early Clinical Assessment of Balance by Gross Motor Function Classification System Level for Children With Cerebral Palsy. Phys Ther [Internet]. 2019 Feb 1 [cited 2025 Apr 14];99(2):217–28. Available from: https://academic.oup.com/ptj/article/99/2/217/5298168

72. Rygelová M, Uchytil J, Torres IE, Janura M. Comparison of spatiotemporal gait parameters and their variability in typically developing children aged 2, 3, and 6 years. Fernandez T, editor. PLOS ONE [Internet]. 2023 May 11 [cited 2025 Sept 1];18(5):e0285558. Available from: https://dx.plos.org/10.1371/journal.pone.0285558

73. Dewar R, Love S, Johnston LM. Exercise interventions improve postural control in children with cerebral palsy: a systematic review. Dev Med Child Neurol [Internet]. 2015 June [cited 2018 Dec 24];57(6):504–20. Available from: http://doi.wiley.com/10.1111/dmcn.12660

74. Curtis DJ, Butler P, Saavedra S, Bencke J, Kallemose T, Sonne-Holm S, et al. The central role of trunk control in the gross motor function of children with cerebral palsy: a retrospective cross-sectional study. Dev Med Child Neurol [Internet]. 2014 Nov [cited 2017 Nov 6];57(4):351–7. Available from: http://doi.wiley.com/10.1111/dmcn.12641

75. Talgeri AJ, Nayak A, Karnad SD, Jain P, Tedla JS, Reddy RS, et al. Effect of Trunk Targeted Interventions on Functional Outcomes in Children with Cerebral Palsy-A Systematic Review. Dev Neurorehabilitation. 2023 Apr 3;26(3):193–205.

76. Curtis DJ, Woollacott M, Bencke J, Lauridsen HB, Saavedra S, Bandholm T, et al. The functional effect of segmental trunk and head control training in moderate-to-severe cerebral palsy: A randomized controlled trial. Dev Neurorehabilitation [Internet]. 2018 Feb 17 [cited 2018 June 6];21(2):91–100. Available from: https://www.tandfonline.com/doi/full/10.1080/17518423.2016.1265603

77. ElBasatiny H, Abdelaziem A. Effect of Trunk Exercises on Trunk control, Balance and Mobility Function in Children with Hemiparetic Cerebral Palsy. Int J Ther Rehabil Res [Internet]. 2015 [cited 2024 July 17];4(5):236. Available from: http://www.scopemed.org/fulltextpdf.php?mno=195778

78. Unger M, Jelsma J, Stark C. Effect of a trunk-targeted intervention using vibration on posture and gait in children with spastic type cerebral palsy: A randomized control trial. Dev Neurorehabilitation [Internet]. 2013 Apr [cited 2015 Nov 24];16(2):79–88. Available from: http://www.tandfonline.com/doi/full/10.3109/17518423.2012.715313

79. Beyaert C, Vasa R, Frykberg GE. Gait post-stroke: Pathophysiology and rehabilitation strategies. Neurophysiol Clin Neurophysiol [Internet]. 2015 Nov [cited 2024 Jan 23];45(4–5):335–55. Available from: https://linkinghub.elsevier.com/retrieve/pii/S0987705315000696

80. Nielsen JB, Christensen MS, Farmer SF, Lorentzen J. Spastic movement disorder: should we forget hyperexcitable stretch reflexes and start talking about inappropriate prediction of sensory consequences of movement? Exp Brain Res [Internet]. 2020 May 7 [cited 2020 May 29]; Available from: http://link.springer.com/10.1007/s00221-020-05792-0

