## Supplementary material for "Investigating rehabilitation by activities involving the trunk to improve balance and gait control in young children with cerebral palsy: a randomized open-label crossover trial protocol": RAIT in CP CPP protocol ENGLISH

**COMMITTEE FOR THE PROTECTION OF PERSONS**

| Designation | Registration No. |
| --- | --- |
|  | 2023-A01969-36 |
| **INSURANCE** |  |
| HDI Global SE | "Tour Trinity, 1 bis place de la Défense", CS 20298, "92035 La Défense, CEDEX" |
|  | Policy No.: 0100534514058 240024 |

**Research Category** RIPH 1 ☐ RIPH 2 ☒ RIPH 3 ☐ *(Note: RIPH refers to "Recherche Impliquant la Personne Humaine," the French legal framework for research involving human subjects. Category 2 pertains to interventional research with minimal risks and constraints.)*

**Sponsor:** **Name:** Institut Régional de Médecine Physique et de Réadaptation Louis PIERQUIN, affiliated with the DEVAH EA3450 research laboratory (development, adaptation, disability) of the University of Lorraine **Address:** 75, boulevard Lobau, CS 34209, 54042 Nancy Cedex, France **Tel:** +33 3 83 52 97 00 **Email:**

**Signatures**

**Sponsor** *[Signature of M. Jonathan PIERRET]* M. Jonathan PIERRET Head of the Research and Innovation Unit Institut Régional de Réadaptation UGECAM Nord-Est 75, Bld Lobau - CS 34209 54042 NANCY CEDEX

**Principal Investigator** *[Signature of Pr Christian Beyaert]* Pr Christian Beyaert

**Sponsor:** **Name:** Institut Régional de Médecine Physique et de Réadaptation Louis PIERQUIN, affiliated with the DEVAH EA3450 research laboratory (development, adaptation, disability) of the University of Lorraine **Address:** 75, boulevard Lobau, CS 34209, 54042 Nancy Cedex, France **Tel:** +33 3 83 52 97 00 **Email:**

**Applicant:** Person authorized by the sponsor to submit the file to the Committee for the Protection of Persons **First Name, Last Name:** Jonathan Pierret **Title:** Doctor of Life and Health Sciences **Position:** Head of the Clinical Research and Innovation Unit **Address:** 75, boulevard Lobau, CS 34209, 54042 Nancy Cedex, France **Tel:** +33 3 83 52 67 61 **Email:**

**Coordinating Investigator:** **First Name, Last Name:** Christian Beyaert **Title:** University Professor-Hospital Practitioner (PU-PH) in Physiology, Pediatrician **RPPS No.:** 100022353976 **Address:** CMPRE, 46 rue du Doyen Parisot – CS 20002, 54630 Flavigny-sur-Moselle **Secretariat Tel:** +33 3 54 59 19 23 **Email:**

**Other investigators, collaborators, and scientists** Annex 1 presents the investigators, collaborating physicians, and scientists involved in this research protocol.

**Clinical Investigation Site** Annex 2 presents the clinical investigation sites.

**Scientific Committee:** The scientific committee is composed of the following individuals:

- **Coordinating Investigator:** Pr Christian Beyaert
- **Investigator(s) specializing in the pathology and therapies studied:** Dr. Fanny Dalmont
- **Sponsor's Representative:** Jonathan Pierret
- **Methodologist(s):** Christelle Requena
- **Data Collection and Management:** Stella Zografou

**History of Protocol Modifications:**

| Version | Date | Description of Modifications |
| --- | --- | --- |

**List of Abbreviations**

- **ANSM:** National Agency for the Safety of Medicines and Health Products (Agence Nationale de Sécurité du Médicament)
- **CNIL:** National Commission on Informatics and Liberty (Commission Nationale Informatique et Liberté)
- **CPP:** Committee for the Protection of Persons (Comité de Protection des Personnes)
- **IRR:** Regional Institute of Rehabilitation (Institut Régional de Réadaptation)
- **MPR:** Physician in Physical and Rehabilitation Medicine (Médecin en Médecine Physique et de Réadaptation)
- **PMSI:** Medical Information Systems Program (Programme de Médicalisation des Systèmes d’Information)
- **RIPH:** Research Involving the Human Person (Recherche Impliquant la Personne Humaine)

**Table of Contents**

- List of Abbreviations
- Table of Contents
- ActivTronc Protocol
  1. Scientific Context
  2. Objective(s), Endpoint(s), and Hypothesis(es) of the Study
  - 2.1. Primary Objective and Hypothesis
  - 2.2. Primary Endpoint
  - 2.3. Secondary Objective(s) and Hypotheses
  - 2.4. Secondary Endpoints
  3. Methodology
  - 3.1. Study Type
  - 3.2. Study Population
    - 3.2.1. Inclusion Criteria
    - 3.2.2. Non-Inclusion Criteria
    - 3.2.3. Sample Size Calculation
  - 3.3. Randomization
  - 3.4. Experimental Plan
  4. Conduct of the Research
  - 4.1. Child Recruitment Procedure
  - 4.2. Rehabilitation Content
  - 4.3. Description of Assessments Performed During the Research
    - 4.3.1. Clinical Assessments
    - 4.3.2. Instrumental Analysis of Gait and Standing
  - 4.4. Expected Results
  - 4.5. Statistical Analysis Plan
  - 4.6. Study Withdrawal and Stopping Criteria
  - 4.7. Duration of the Research and Compensation
  - 4.8. Exclusion and Exclusivity Period
  5. Benefits and Risks
  - 5.1. Benefits for the Participant
  - 5.2. Risks and Constraints for the Participant
  6. Safety Evaluation
  - 6.1. Adverse Events
    - 6.1.1. Definitions
    - 6.1.2. Serious Adverse Event(s)
  - 6.2. Procedures to Follow
    - 6.2.1. Procedures to Follow in Case of a Serious Adverse Event
  7. Data Management
  - 7.1. Right of Access to Data
  - 7.2. Source Data
  - 7.3. Participant Anonymity
  - 7.4. Confidentiality
  - 7.5. Archiving
  - 7.6. Rules for Publication of the Final Report
  8. Quality Assurance
  - 8.1. Investigator's Commitment - Good Clinical Practices
  - 8.2. Data Quality Control
  - 8.3. Audit and Inspection
  9. Study Organization
  - 9.1. Scientific Committee
  10. Ethical and Regulatory Considerations
  - 10.1. Research Site Authorization
  - 10.2. Ethical Conduct of the Study
  - 10.3. Committee for the Protection of Persons
  - 10.4. Investigator's Responsibilities
  - 10.5. Sponsor's Responsibilities
  - 10.6. Data File Declaration
  11. Bibliography
  12. Annexes
  - 12.1. Annex 1: List of Investigators and Scientific Collaborators
  - 12.2. Annex 2 : List of Clinical Investigation Sites

**ActivTronc Protocol**

**1. Scientific Context**

Cerebral palsy (CP) describes a group of permanent motor and postural disorders that limit activity, caused by lesions to the developing brain in utero or post-partum. Subsequent reorganizations of central nervous system functions in children with CP also lead to motor disorders (1). The typical development of segmental trunk control, which occurs progressively during childhood (2), is closely linked to gross motor performance in young infants (3) and is significantly delayed in premature infants (4). From early infancy, children with CP exhibit axial control disorders and abnormal postural control in a sitting position (5). We recently showed that children with CP had specific impairments in trunk control during a self-stabilization task on an unstable sitting device compared to typically developing (TD) children (6). These deficits in axial postural control (and particularly in trunk control) remain present throughout the motor development period—even after the child can stand and walk independently (7).

During walking, children with CP show significant deviations in trunk kinematics and kinetics; the ranges of motion of the trunk in all three planes of space are much larger than in TD children (8). These deviations are associated with poor dynamic balance control during walking (9), resulting in wider step widths (10), greater variability in step length, greater accelerations of the head, thorax (upper trunk), lower back (L3 region), pelvis, and the body's center of mass in all three planes throughout the gait cycle, and greater lower back instability compared to TD children (11,12). Since the trunk and lower limbs interact reciprocally during walking, in children with CP, trunk deviations lead to lower limb deviations, and vice versa (7,13).

Toe walking is one of the most common lower limb deviations in children with CP. It is defined as the absence of the first rocker (heel rocker) when children touch the ground with a flat foot or with the forefoot at initial contact (IC) (14). Thus, IC is immediately followed by the second rocker, as the tibia rolls over the ankle. At IC, the ankle angle is often in plantar flexion, i.e., in equinus. The absence of the first rocker in children with CP is also associated with intense and early energy absorption and negative work done at the ankle joint (15), which decelerates ankle dorsiflexion and the anterior tilt of the tibia during the weight acceptance (WA) phase of walking (defined as the initial period of combined energy absorption at the lower limb joints) (16). This behavior involves early activity of the triceps surae, which begins in late swing and lasts throughout the WA phase (17). It is generally believed that prolonged activity of the plantar flexors during walking in children with CP is due to spasticity (hyperexcitable stretch reflexes) and induces equinus (1). Nevertheless, the existence and/or functional significance of exaggerated stretch reflexes during walking in spastic patients has been debated for decades (18). In particular, it is unlikely that spasticity contributes to toe walking in children with CP; soleus activity during the swing phase is reduced (15), and exaggerated reflex activity is absent (19). In contrast, toe walking in children with CP and in typically developing (TD) children is characterized by feedforward control of the ankle muscles—suggesting that this walking pattern is part of an adaptive process (20).

When the second rocker begins, the plantar flexors decelerate ankle dorsiflexion, slow the forward progression of the trunk, and support the body by accelerating it upward (21). This upward acceleration of the body can be achieved either by moving the center of mass (CoM) upward or by decelerating the downward movement of the CoM. In children with CP, the activity of the plantar flexors associated with the early second rocker in WA results in negative ankle power, which slows ankle dorsiflexion and thus decelerates the body's CoM downward and forward (22). All these actions contribute to the greater negative work done on the CoM by the leading leg in children with CP compared to TD children (23). The presence of the aforementioned trunk control disorders and impaired dynamic balance control during walking in children with CP suggests that the early activation of plantar flexors and the high negative ankle power during WA associated with toe walking could correspond to an adaptive mechanism to decelerate the forward and downward displacements of the trunk, in order to compensate for poor balance and poor postural control of the trunk.

In children with cerebral palsy (CP), aged 5 to 12 years, with impaired trunk control and independent toe walking, we recently showed, compared to TD children, a greater step width (due to impaired balance) (10), a greater peak of anterior sternum deceleration, downward sacrum deceleration, and negative ankle power during the WA phase (24) **(Figure 1).**

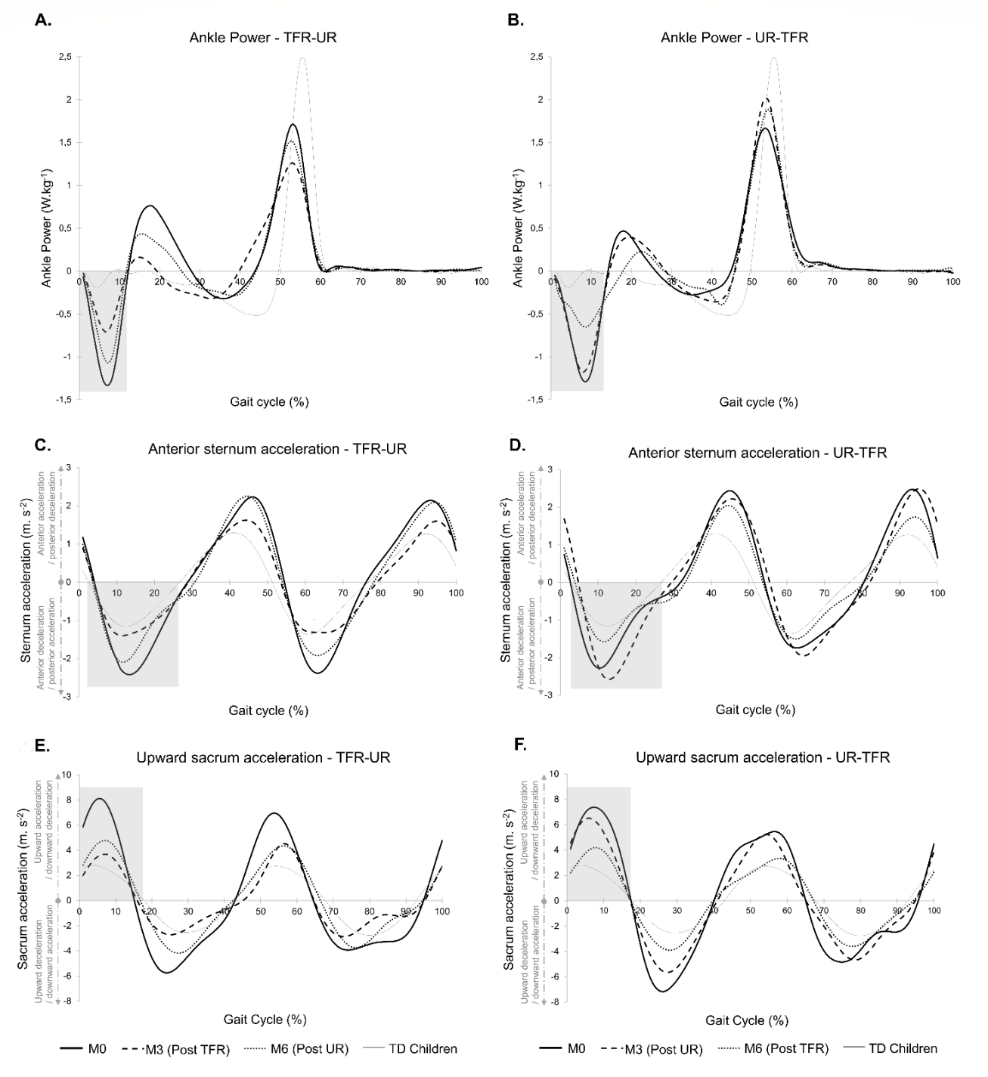

**Figure 1:** Mean total ankle power (A, B), mean anteroposterior acceleration of the sternum marker (C, D), and mean upward acceleration (i.e., downward deceleration) of the sacrum marker (E, F), during walking, in children with cerebral palsy (CP) who performed TFR then UR (A, C, E, n=8) or UR then TFR (B, D, F, n=9), and in TD children (all panels, n=17). TFR: trunk-focused rehabilitation (=RAIT); UR: usual rehabilitation (=RH); TD: typically developing. The black lines correspond to the CP group (solid line for M0, dotted line for M3, and dashed line for M6) and the solid gray line corresponds to the TD group at M0. The rectangle corresponds to the area of interest for the peak during the weight acceptance phase.

The peak deceleration of the sternum and sacrum were significantly correlated with the peak negative ankle power during the WA phase, which is consistent with the action of the plantar flexors on the trunk or the CoM (22,25). This cross-over study also allowed for the evaluation of the effect of a rehabilitation based on activities strongly involving the trunk (RAIT) for 3 months compared to a usual rehabilitation (RH) mainly based on conventional rehabilitation targeting muscle groups (stretching, strengthening, tone reduction) of the lower limbs. Only RAIT significantly improved trunk postural control, as evidenced by the improvement in the Trunk Control Measurement Scale score (26) and postural self-stabilization on an unstable sitting device (6). Furthermore, only RAIT significantly reduced the peaks of sternal and sacral deceleration and the coupled negative ankle power in the WA phase (see Figure 1). It is thus strongly suggested that the plantar flexors compensate for impaired trunk postural control—a key target of rehabilitation in children with CP.

Therefore, there is a strong interest in rehabilitating the trunk in children with CP. Indeed, while our study shows benefits on parameters related to dynamic balance during walking, many other studies have made the trunk the main target of their intervention (27–32). These studies notably show an improvement in trunk control, but also in walking (spatio-temporal parameters, performance on the Timed-Up-and-Go, the 1-minute walk test, and the 6-minute walk test) as well as gross motor function. Moreover, studies that have tested hippotherapy protocols in children with CP link the observed improvements in balance, walking, and gross motor function with the improvements induced in trunk control (33–36).

Due to the importance of trunk control on the gross motor function, balance, and walking of children with CP, and given all the studies showing the benefit of a trunk-focused rehabilitation, trunk-focused rehabilitation protocols are now prescribed to patients in our center whose profile seems to indicate an interest in trunk strengthening. The study of the effect of RAIT on the ability to stabilize in an unstable sitting position and on the dynamics of the trunk and ankle during walking, which we conducted, was carried out in children with CP aged 5 to 12 years, partly due to technical limitations related to the unstable seat and the three-dimensional motion analysis with many markers to be placed on the skin, which are difficult to perform on children under 5 years of age. However, it seems relevant to carry out a rehabilitation like RAIT early, from the age of 18 months, to improve postural balance control and walking dynamics in children with CP.

With this in mind and to simplify the functional exploration of walking dynamics, and to be able to evaluate the effects of the RAIT prescribed to our youngest patients, we have acquired inertial measurement units to measure the deceleration of the sternum and the sacrum (L5) and a pressure-sensitive walkway (**Figure 2**) to analyze gait instability via spatio-temporal parameters and to measure, at the beginning of the stance phase, the extent of the early forward shift of the plantar pressure associated with the toe-walking pattern. The analysis of postural control of standing is also simplified through the use of an inertial measurement unit placed at L5 to estimate the displacements of the body's center of mass (mSway software). Clinical scores are also used for an assessment of balance and global functional motor skills. Furthermore, RAIT has other potential benefits such as an improvement in the functional use of the upper limb and hand, observed on the sidelines of the study in children aged 5 to 12 years (24). Also, in this study we explore the motor function of the upper limb and hand via a questionnaire addressed to the parents.

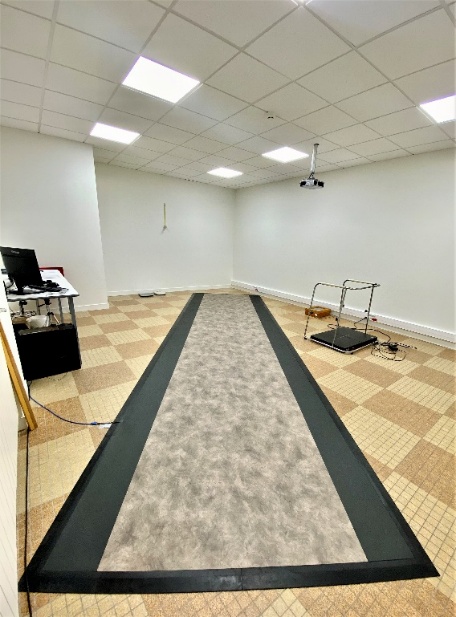

**Figure 2:** Walkway equipped with relative pressure sensors (Zeno®) allowing for an extended and simplified analysis of spatio-temporal parameters and the dynamics of plantar footprints

**2. Objective(s), Endpoint(s), and Hypothesis(es) of the Study**

**2.1. Primary Objective and Hypothesis**

The primary objective of the study is to show that the RAIT protocol, prescribed over a 3-month period, and not a usual rehabilitation (RH), reduces the peak forward deceleration of the sternum at the beginning of the stance phase (the first 25% of the gait cycle) in children with CP aged 18 months to 5 years and 6 months who walk independently or with intermittent use of a walking aid (walker, crutches, etc.), which corresponds to levels I to II of the Gross Motor Function Classification System (GMFCS) (37). Children who intermittently use a walking aid must be able to walk independently for a distance of about 10 m to allow for assessment on the walkway.

**2.2. Primary Endpoint**

Peak forward deceleration of the sternum during the first 25% of the gait cycle.

**2.3. Secondary Objective(s) and Hypotheses**

The secondary objectives, for the secondary endpoints described in paragraph 2.4 below, aim to: (1) show an impairment of these criteria in the study's children with CP compared to typically developing (TD) children; (2) show a significantly greater improvement in these criteria after 3 months of RAIT than after 3 months of RH; (3) show a further improvement in these criteria when the duration of RAIT is increased.

**2.4. Secondary Endpoints**

(1) Variables related to the quality of spontaneous walking:

- Peak downward deceleration of L5 during the first 25% of the gait cycle
- Percentage of load on the anterior half of the foot during the first double support phase
- Step width normalized to pelvis width
- eGVI: enhanced Gait Variability Index (38,39)

(2) Variables related to postural control of standing:

- Mean velocity of horizontal displacement of the body's center of mass
- 95% confidence ellipse of the area of horizontal displacement of the body's center of mass

(3) Functional variables assessed by clinical scores:

- Score on the Early Clinical Assessment of Balance (ECAB) (40)
- Score on the Gross Motor Function Measure 66 Item Set (GMFM 66 IS) (41)
- Edinburgh Visual Gait Score (42)

**3. Methodology**

**3.1. Study Type**

Single-center randomized controlled interventional study.

**3.2. Study Population**

Children with cerebral palsy presenting with gait pattern disorders seen in outpatient consultation and/or treated (day or full hospitalization) at the CMPRE of Flavigny-sur-Moselle. Cerebral palsy is a long-term condition requiring well-codified specialized medical follow-up. In the Lorraine region, the multidisciplinary therapeutic follow-up of this population is coordinated by a network of physicians specializing in physical and rehabilitation medicine, neuro-pediatrics, and pediatrics working in several institutions. The children with CP recruited for this study will be children with spastic hemiplegia or diplegia, with independent walking without a walking aid (GMFCS I to II). In the case where the child has diplegia, only the lower limb with the most spastic soleus will be analyzed. The most spastic soleus is defined as the one for which the greatest difference is observed between the range of slow passive dorsiflexion movement and the angle obtained using the maximum dorsiflexion speed V3, according to the Modified Tardieu Scale (43).

**3.2.1. Inclusion Criteria**

**For children with CP:**

- Age between 18 months and 5 years and 6 months
- CP of spastic diplegia or spastic hemiplegia type, GMFCS I to II
- No or moderate contracture of the triceps surae (ankle dorsiflexion: > 5° during clinical examination, knee extended)
- Sufficient level of understanding to perform the activities involving the trunk in the form of self-exercises (rehabilitation protocol) as well as the clinical assessments and functional explorations.
- Acceptance of the physical therapist in charge of the child's follow-up to collaborate in the implementation of the RAIT
- Affiliated with a social security scheme

**For TD children:**

- Age between 18 months and 5 years and 6 months
- Walking acquired before the age of 18 months
- Sufficient level of understanding to perform the clinical assessments and functional explorations
- Affiliated with a social security scheme

**3.2.2. Non-Inclusion Criteria**

**For children with CP:**

- Previous lower limb surgery within the last year
- Botulinum toxin A injection within the last 6 months
- Any change in rehabilitative and/or orthopedic management within the last 2 months
- Hip flexion contracture > 20°
- Presence of subacute or chronic pain during standing or walking

**For TD children:**

- Neurological and/or orthopedic disorder that could influence walking

**3.2.3. Sample Size Calculation**

The calculation of the number of subjects needed is based on the primary endpoint. The distribution of the peak sternum deceleration at the beginning of the stance phase was Gaussian in our previous study. On average, this distribution was centered on 3.0 ± 1.1 m.s-2 (mean ± 1 standard deviation) in children with CP and on 1.1 ± 0.3 m.s-2 in TD children (24). The expected effect of RAIT is a minimum reduction in the peak sternum deceleration at the beginning of the stance phase of one-third of its value, similar to the effect of RAIT in the previous study (24). To demonstrate an effect of RAIT using a repeated measures ANOVA in which we expect a small effect size, with an alpha level set at 0.05 and a power of 80%, a total sample size of 24 patients, i.e., 12 per group, is necessary (calculation performed using G*Power 3.1 software; **Figure 3).** We expect greater variability in dynamic gait variables (expected in children aged 18 months to 5 years and 6 months compared to children aged 5 to 12 years). Therefore, we plan to recruit 32 children with CP (16 per group), to account for this greater variability and to compensate for potential dropouts.

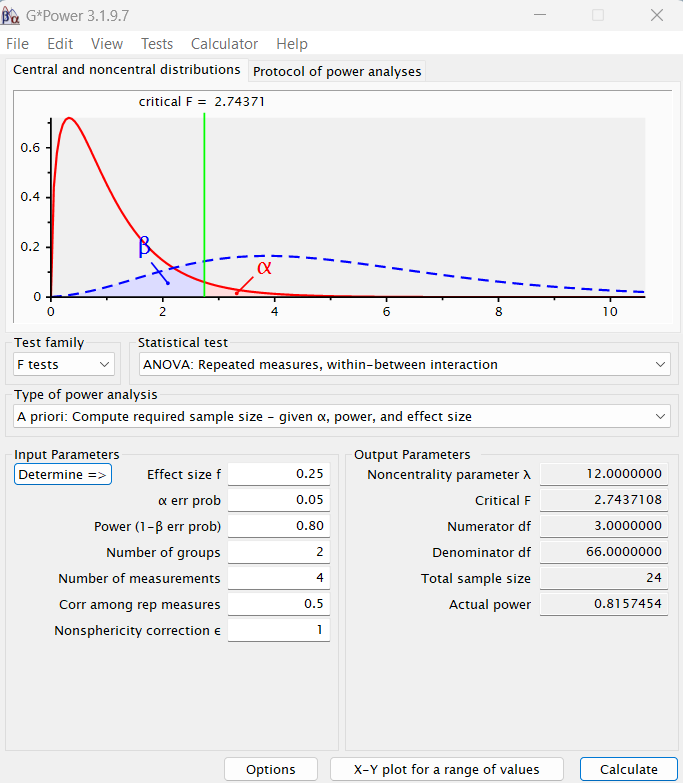

**Figure 3:** Calculation of the number of subjects needed, performed using G*Power 3.1 software.

**3.3. Randomization**

The sample of children with CP will be divided into two groups: PC1 and PC2, with individual random assignment of participants to each group in blocks of 4 or 8 children. This assignment will be carried out using a specific program coded in Matlab software. A control sample of TD children (n=32) will be matched for age and sex with the children with CP.

**3.4. Experimental Plan**

The project includes a clinical examination and instrumental functional explorations at four time points. The experimental plan is shown in Figure 4. The assessments are described in chapter "4.3. Measurements – Description of assessments performed during the research." Assessments will be conducted at M0, M3 (M0 + 3 months), M6 (M0 + 6 months), and M12 (M0 + 12 months) for children with CP and at M0 for TD children. Thus, three rehabilitation periods will be planned: the first between M0 and M3, the second between M3 and M6, and the third between M6 and M12. A first group of children (group PC1) will continue their usual rehabilitation (RH) for the first 3 months and then will have RAIT for the following 9 months (PC1 = RH-RAIT). The second group of children, PC2, will immediately have RAIT for the 12 months of the study (PC2 = RAIT-RAIT). As the previous study conducted in children with CP aged 5 to 12 years showed a therapeutic effect only related to RAIT (and not to RH), RAIT will be continued for the children who benefited from it from the first 3-month period and for all children for the last 9 months of the study. Such a plan will allow to: 1) ensure that all children with CP benefit from RAIT; 2) measure the effect of RAIT over 3 months compared to that of RH; 3) test the reproducibility of the effect of RAIT over 3 months between the two groups and 4) test the effect of an additional 3 months of RAIT in the PC2 group (M6 versus M3) and 6 months in both groups (M12 versus M6) (**Figure 4**).

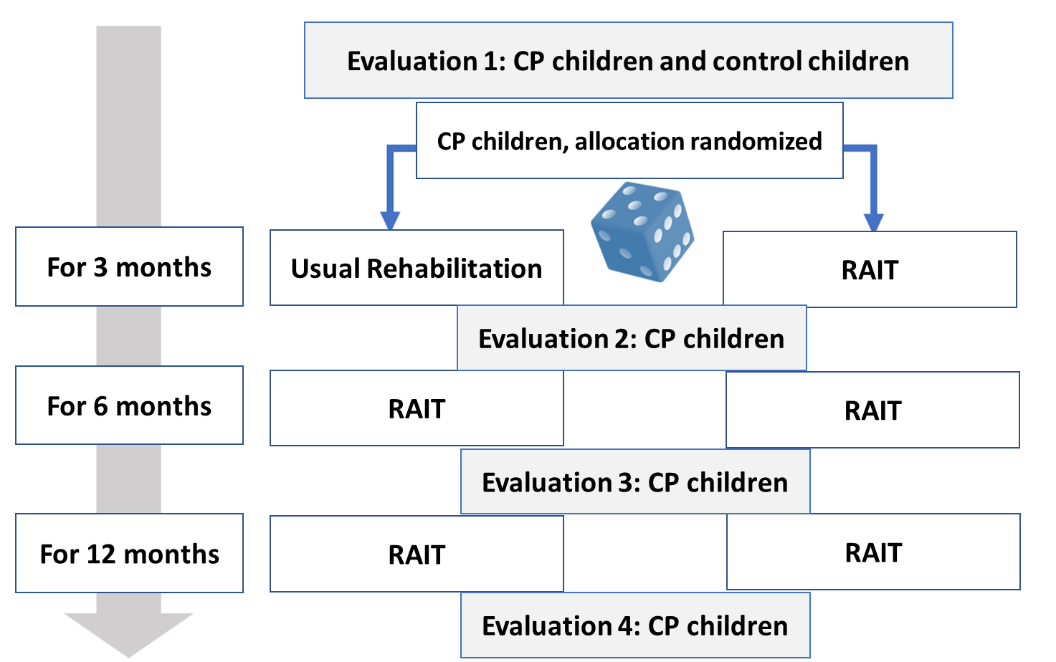

**Figure 4:** Experimental plan of the study. RAIT: rehabilitation through postural activities involving the trunk; conventional rehabilitation is based on the treatment of muscle groups (stretching, strengthening, tone reduction) of the lower limbs

**4. Conduct of the Research**

**4.1. Child Recruitment Procedure**

The Center for Physical Medicine and Rehabilitation for Children (CMPRE) in Flavigny-sur-Moselle is a regional center providing various functional assessment and treatment services for children with orthopedic, neurological, or neuro-orthopedic abnormalities, including children with CP. Functional motor assessment sessions are offered and include clinical assessments (neuro-orthopedic evaluations and functional motor scores) and functional explorations of walking and standing using a pressure-sensitive walkway, video cameras, and inertial measurement units.

The initial diagnosis and medical follow-up of children with CP are carried out by physicians interacting in different settings: RAFAEL network (Lorraine Family Support Network), neuro-pediatric consultations at the Brabois Children's Hospital in Vandœuvre-lès-Nancy, Early Medico-Social Action Centers (CAMSP) in Lorraine, private pediatric consultations, or consultations at the Center for Physical Medicine and Rehabilitation for Children in Flavigny-sur-Moselle. The solicitation of these different physicians across Lorraine should allow for the inclusion of 32 children with CP over a 30-month period.

When one of these physicians, during a consultation with a child with CP, prescribes a functional motor assessment associated with a medical consultation at the CMPRE and this child is potentially eligible for the study, they will orally inform the parents in an interactive exchange and give them the information sheet and consent form in preparation for a possible inclusion in the study, if they agree. When scheduling the appointment at the CMPRE, the contact information of the physical therapist who treats the child will be collected so that he or she can be informed about the study and their consent to participate can be obtained, in case of the child's inclusion.

During the medical consultation associated with the child's functional motor assessment, one of the recruiting physicians from the CMPRE (Dr. Christian Beyaert, Dr. Fanny Dalmont, Dr. Marion Birck) will explain the study to the parents and the child, according to their level of understanding, based on the documents in their possession during an interactive exchange and will offer them to participate if the eligibility criteria are met. If they agree, they will sign the previously provided consent form, and their child's inclusion in the group with initial RAIT or in the group with initial usual rehabilitation will be allocated according to a prior randomization. If the parents wish to think further before accepting or refusing their child's participation in the study, an additional period of 7 days will be offered.

For the control group of TD children, recruitment will be done through an email announcement presenting the study and including the information and consent form, addressed to people in the Greater Nancy Metropolitan Area, within the staff of the University of Lorraine not hierarchically linked to the study investigators, or whose children attend sports and educational associations. The typically developing siblings of the children with CP participating in the study will also be solicited. Like the children with CP, they will be seen in consultation with an investigator physician. During this consultation, the investigator will be responsible for informing the participant and their parents about the nature of the study, its objectives, its methodology, its duration, the expected benefits, the foreseeable constraints and risks including in case of stopping the study before its term, and the medical care arrangements planned at the end of the research if necessary. This informative discussion will be based on the information letter previously provided to the parents.

The parents (and the participating child, within the limits of their young age) will have every opportunity to ask any questions concerning the research and will be informed of their right to refuse to participate in the research or to withdraw their consent at any time without prejudice and without having to justify the reasons for their decision. The children and their parents will be informed orally in understandable language about the course of the study and the examinations to be performed. The informed consent form will be signed in two copies by the different parties, after an additional reflection period if the parents so desire. The investigator and the parents of the child participating in the research will each keep an original copy of the consent form. The information for parents and any other written information given to them must be updated if new data becomes available that could change their opinion or if an amendment to the protocol requires a correction of the information and consent form. The investigator will inform the child's parents of these changes and ask them to confirm their agreement by dating and signing a new consent form. Any modification of these documents must be approved by the CPP.

**4.2. Rehabilitation Content**

Each child with CP will continue to participate in one to three rehabilitation sessions per week for a total of 12 months (initially 3 months of URor RAIT, then 9 months of RAIT) under the direction of the child's physical therapist, provided that he or she agrees to participate in the team and has not participated in the previous study using RAIT. The physical therapist is then initially informed of the principles of RAIT and receives detailed instructions on its content shortly before the start of the RAIT treatment period. For children who are treated at the CMPRE in Flavigny-sur-Moselle, a physical therapist from the center will be responsible for monitoring the patient's rehabilitation. Each session lasts about 30 to 45 minutes, and the child must receive exercises to be performed daily at home for 15 to 30 minutes, depending on the parents' availability.

Usual Rehabilitation corresponds to the type of rehabilitation the child was already receiving before the study; most often, it combines in varying degrees muscle stretching and strengthening, muscle tone reduction (e.g., neurodevelopmental treatment according to the Bobath concept), and therapist-facilitated training of upper and lower limb motor skills. These therapies, involving limited groups of muscles in stretches or elementary actions, usually target the muscles of the lower limbs, sometimes those of the upper limbs, and more rarely those of the trunk. During the study's UR period for the PC1 group, the physical therapist will propose to the parents, according to their availability and ability, to perform a suitable selection of the stretches and elementary actions that he or she performs in the office on the child daily for 15 to 30 minutes.

The RAIT program is not based on strengthening elementary trunk muscles, but on improving postural control and balance of the whole body, including the trunk and other affected muscles, by means of actions performed autonomously by the child in intermediate postures involving the trunk (24). Thus, the principle of this approach is, during autonomous actions in intermediate postures, to exploit the fundamental automatic control of postural support and balance to improve the use of affected muscles in support and balance not only during these actions but also during all postural and locomotor tasks, an original approach proposed for the rehabilitation of stroke patients (44). The child must control their balance during various voluntary actions, from intermediate postures such as repeatedly alternating between the four-point kneeling posture and the so-called "cobra" posture, or swinging the body back and forth from the so-called "downward-facing dog" posture. These autonomous actions are less difficult than standing and walking, but beneficial effects on the latter are expected. From intermediate postures, the child also performs more difficult trunk movements, requiring the dissociation of the movements of the shoulder and pelvic girdles or a reduction of lumbar lordosis. The upper and lower limb rehabilitation goals defined before the study and followed during UR (e.g., hamstring stretches, plantar flexor stretches, and wrist extension) are included in some of the RAIT activities. For example, from a four-point kneeling position with wrists in extension, the child lifts the knees, which achieves hamstring and plantar flexor stretches while engaging the trunk and the whole body to manage balance. Finally, each child receives a selection of activities to be performed daily at home.

**4.3. Description of Assessments Performed During the Research**

A motor function assessment will be carried out at the CMPRE at the beginning of the study and 3, 6, and 12 months later to evaluate the effect of the treatment. This assessment includes clinical evaluations and an instrumental evaluation of walking and standing.

**4.3.1. Clinical Assessments**

Several clinical assessments will be performed by a physical therapist:

**Neuro-orthopedic evaluation** A therapist will perform a neuro-orthopedic evaluation of the lower and upper limbs. This examination consists of measuring, (i) using a goniometer, the ranges of motion of the main joints in one or more planes, (ii) the muscle strength of the major muscle groups by asking the subject to move a joint against resistance, and (iii) the spasticity of the main muscles, by moving a joint at a slow then a fast speed to elicit a stretch reflex. This evaluation lasts about 45 minutes.

**GMFM-66-IS** The Gross Motor Function Measure 66 (GMFM-66) is a standardized clinical score with 66 items that assesses the gross motor function and its evolution over time in children with cerebral palsy (45). The GMFM-66-IS is a faster (about 20 to 30 minutes versus 60 to 80 minutes) and validated scoring method for the GMFM-66, using 15 to 39 items (41,46).

**The Early Clinical Assessment of Balance (ECAB)** This is a 13-item clinical scale that assesses postural stability (balance ability) in children with cerebral palsy, with two subscales: one dedicated to head and trunk postural control, and one dedicated to sitting and standing postural control. This scale is validated for children from 1.5 to 11 years old, regardless of the GMFCS level (40,47). The optimal score is 100. The administration of this scale takes about 15 minutes.

**Gross Motor Function Classification System (GMFCS) - Family Report Questionnaire** This questionnaire for parents, based on the child's voluntary movements for sitting, transfers, and mobility, helps to classify the severity of the child's cerebral palsy into one of 5 levels. The questionnaire takes about 2 minutes to complete.

**"Reaching Hand" Questionnaire** This is a questionnaire for parents to assess the abilities of the upper limb and hand in different functional situations for their child with CP (2023 Ma). This questionnaire, validated for children with CP from the age of 2, will assess the expected improvement in hand and upper limb functions related to RAIT. The questionnaire takes about 15 minutes to complete.

**4.3.2. Instrumental Analysis of Gait and Standing**

The Zeno gait analysis walkway (Figure 2) is a mat made of pressure sensors that collects spatio-temporal gait parameters and the progression of foot pressure patterns. These parameters are commonly used to analyze locomotion, identify gait disorders, and evaluate the effect of therapeutic interventions. The patient is asked to walk back and forth on the walkway.

Four inertial measurement units (**Figure 5**) will be used in addition to the walkway, placed on the skin with a double-sided hypoallergenic adhesive: one on the sternum, one over the 5th lumbar vertebra (L5), and one on each foot. This will allow for the measurement, at the beginning of the stance phase, of the peak decelerations of the upper part of the trunk (sternum) and the lower part of the trunk near the center of mass (L5), and to measure the transverse orientation of the feet during walking.

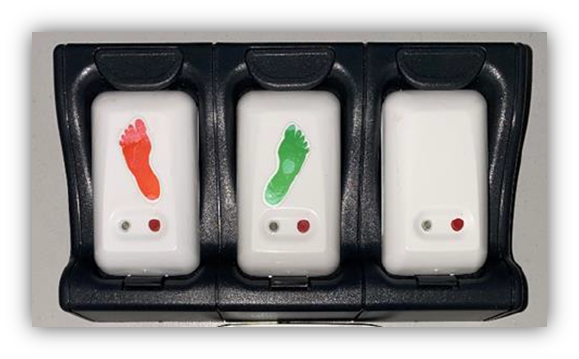

**Figure 5:** Example of the mTest3 inertial measurement units (mHealth Technology) used.

Two video cameras, one in profile and one from the front, will allow for the analysis of gait kinematics using the Edinburgh Visual Gait Score, which is validated for children with cerebral palsy (42). The analysis of walking at a self-selected speed, carried out in about 10 minutes, will allow us to collect the following variables:

- **The peak anterior deceleration of the sternum at the beginning of the stance phase** (the first 25% of the gait cycle), which we have shown to be reduced after RAIT in children with CP aged 5 to 12 years (24), will constitute the primary endpoint.
- **The peak downward deceleration of L5 at the beginning of the stance phase**, which we have shown to be reduced after RAIT in children with CP aged 5 to 12 years (24), complements the primary variable for the dynamic behavior of the trunk during walking.
- **The enhanced Gait Variability Index (eGVI)**, recently published, is a composite score based on 9 spatio-temporal parameters that quantifies the distance between the amount of variability observed in an asymptomatic reference group and the amount of variability observed in the patient (38,39). This index assesses instability during walking and the risk of falling. It is usually high in children with CP (48,49).
- **The step width** (between the 2 feet), which is high in children with CP, is one of the strategies to reduce the risk of falling in case of unstable walking (50).
- **The anterior foot load during the 1st double support phase** is a variable obtained on a pressure-sensitive walkway, defined by the ratio between the integrated pressure of the anterior half-foot and the integrated pressure of the entire foot during the 1st double support phase. This variable will be higher when the foot makes contact with the ground on the forefoot, as in children with CP, and lower when the foot makes contact on the heel, as in TD children. This variable, developed for this study, is related to the early anterior displacement of the center of pressure frequently observed in children with CP (51).
- **The Edinburgh Visual Gait Score** (Read 2003) is a score developed for children with cerebral palsy, assessing the extent of kinematic deviations compared to typically developing children.

The analysis of standing will be performed by asking the child to stand still for 30 seconds, 3 times. The analysis of the displacement of the inertial measurement unit placed at L5 using the mSway software from mHealth will allow for the estimation of the mean velocity and the 95% confidence ellipse of the area of horizontal displacement of the body's center of mass (ref).

**4.4. Expected Results**

**For children with CP compared to TD children, it is expected**:

- A higher value for the peak forward deceleration of the sternum and downward deceleration of L5 at the beginning of the stance phase.
- Higher values for step width and the eGVI.
- A greater anterior foot load during the 1st double-support phase.
- Higher values for the mean velocity and area of displacement of the center of mass.
- A higher value for the Edinburgh Visual Gait Score.
- Lower scores on the ECPE, the GMFM-66-IS, and the "Reaching Hand" questionnaire.

**In children with CP, the expected effect after RAIT is an improvement in the endpoints**:

- Decrease in the peak forward deceleration of the sternum and downward deceleration of L5 at the beginning of the stance phase.
- Decrease in step width, the eGVI, and the anterior foot load during the 1st double-support phase.
- Decrease in the Edinburgh Visual Gait Score.
- Decrease in the mean velocity and area of displacement of the center of mass.
- Increase in the scores of the ECPE, the GMFM-66-IS, and the "Reaching Hand" questionnaire.

**4.5. Statistical Analysis Plan**

Statistical analyses will be performed using general linear models, as the overall sample size (N=64) and the size of each group (n=32) are considered sufficient for applying such procedures. The conditions for using these tests will be verified beforehand, particularly the normal distribution of residuals (using quantile-quantile plots and Shapiro-Wilk tests) and the homogeneity of variance (Levene's tests). If the data do not meet these conditions, non-parametric equivalents will be used.

In the first phase, the group effect (Children with CP vs. TD Children) will be analyzed using multivariate analysis of variance (MANOVA) procedures on the dependent kinematic (step width), dynamic (sternum and L5 decelerations and anterior foot position during the 1st double-support phase), and clinical (ECAB, GMFM-66-IS, and "Reaching Hand" questionnaire) variables. A multiple linear regression analysis will also be performed, with the anterior foot position during the 1st double-support phase as the dependent variable and the sternum and L5 decelerations at the beginning of the stance phase as the regressors.

Furthermore, the effects of the specific rehabilitation (2 Groups x 4 Periods) will be tested using the same type of multivariate procedures (MANOVA) and a 2 Groups [PC1 vs. PC2] × 4 Periods [M0, M3, M6, M12] mixed design, with a particular focus on the Group × Period interaction. If this interaction is significant, the analysis of local comparisons using post-hoc procedures (Tukey's tests) will allow us to examine: 1) the significance of the rehabilitation (significant difference between the two groups at M3 with a reduction in the peak sternum deceleration at the beginning of the stance phase for the group that underwent RAIT between M0 and M3); 2) the reproducibility of the RAIT effect (similar differences between M3 and M6 for the RH-RAIT group and between M0 and M3 for the RAIT-RAIT group); and 3) the enhancement of the RAIT effect when it is continued (between M6 and M0 versus between M3 and M0, and between M12 and M0 versus between M6 and M0 for the RAIT-RAIT group; and between M12 and M3 versus between M6 and M3 for the RH-RAIT group).

**4.6. Study Withdrawal and Stopping Criteria**

The parents of the participating child are free to withdraw their child from the study at any time without needing to provide a reason. The protocol will also be terminated if a socio-medical imperative arises during the study that requires it to be stopped. In such a situation, the final assessment will be performed at the time of study discontinuation, if possible. The study will be stopped after the last visit of the last enrolled patient. The sponsor can decide to terminate the study prematurely. The sponsor must then inform the CPP within 15 days and state the reasons for this decision. At the end of the study, the sponsor will inform the CPP and the investigating centers within 90 days. In the event of premature study termination, the investigators must promptly inform the participating patients and ensure they receive appropriate follow-up care.

**4.7. Duration of the Research and Compensation**

- **Duration of patient participation in the protocol:** 12 months
- **Duration of the inclusion period:** 30 months
- **Anticipated duration of the research (inclusion + analysis):** 54 months
- No compensation is planned.

**4.8. Exclusion and Exclusivity Period**

- **Exclusion period:** For the entire duration of the study, the participant in this study cannot participate in any other research involving human subjects.
- **Exclusivity:** The participant cannot take part in any other interventional research during this study.

**5. Benefits and Risks**

**5.1. Benefits for the Participant**

All children with CP participating in this study will benefit from RAIT rehabilitation for either 9 or 12 months. If RAIT shows the same effects as in our previous study on children aged 5 to 12, the expected direct benefits are better balance control during sitting and standing and improved walking dynamics (trunk decelerations and foot dynamics). Furthermore, in cases of upper limb motor abnormalities, improved use of the upper limb and hand (associated extension of the elbow, wrist, and fingers; manual dexterity) is also expected, based on unpublished clinical experience. The physical therapists and the patients' parents will be able to continue this care even after the study ends.

**5.2. Risks and Constraints for the Participant**

There are no major risks for the participant concerning the treatments and assessments performed. The postural activities, in which muscles and tendons are stretched, could initially be painful. However, any muscle and tendon pain, should it occur, would be mild and temporary because the stretching of muscles and tendons will be progressive, especially since the child performs the postural activities themselves.

The constraints related to the study are quite significant. The RAIT postural activities or, where applicable, a suitable selection of exercises related to usual rehabilitation prescribed by the physical therapist, must be performed regularly, every day, for at least 15 to 30 minutes (in one or more sessions). The home sessions are complementary to those performed with the physical therapist. An adult will need to be present during home sessions to supervise the child. The postural activities or usual rehabilitation exercises performed by the child at home will be recorded in a logbook so that their progress can be monitored by the physical therapist and the CMPRE team. There will be a total of 4 sessions at the CMPRE: at the start of the study and then at 3, 6, and 12 months. Each session will last several hours (2 to 4 hours) spread over one day and will include the motor function assessment and a medical consultation to review the postural activities. A member of the CMPRE team will contact the parents via teleconsultation once or twice between each motor function assessment at the CMPRE to answer their questions.

**6. Safety Evaluation**

**6.1. Adverse Events**

**6.1.1. Definitions**

- **Adverse Event:** Any harmful occurrence in a person participating in research involving human subjects, whether or not it is related to the research or the product being studied.
- **Adverse Effect:** An adverse effect of a research study corresponds to any adverse event caused by the research.
- **New Event/Finding:** Any new data that could lead to a re-evaluation of the benefit-risk ratio of the research or the product under investigation, to changes in the use of this product, in the conduct of the research, or in the research documents, or to the suspension, interruption, or modification of the research protocol or similar research.

**6.1.2. Serious Adverse Event(s)**

In this study, regardless of the intensity or duration of the treatment, any adverse event or effect will be considered serious (SAE) if it:

- Results in death.
- Is life-threatening to the participant.
- Requires hospitalization or prolongation of existing hospitalization.
- Results in persistent or significant disability or incapacity.
- Results in a congenital anomaly or birth defect.
- Constitutes a medically significant situation.

In this protocol, the following will not be considered serious (SAE):

- An event leading to a temporary visit to a hospital consultation, an emergency room, or a day hospital.
- Hospitalizations for elective procedures not associated with a worsening of the clinical condition and not related to the objective of the clinical study, occurring during the clinical study (e.g., cosmetic surgery).
- Hospitalizations (more than one night) or extensions of hospitalization for the following reasons : planned hospitalizations for routine interventions or treatments as part of a pre-defined monitoring or therapy program ; hospitalizations for investigations not related to a change in the participant's condition ; or hospitalization for convenience or social reasons.

**6.2. Procedures to Follow**

All adverse events occurring during the study, whether observed by the physicians or reported by the patients and regardless of the suspected causality, must be recorded in the case report form and followed up appropriately. For each adverse event, the following should be reported whenever possible:

- Its description or diagnosis.
- Its duration (start and end dates).
- Its severity (mild, moderate, severe).
- Its relationship to the investigational product (related/unrelated).
- The measure(s) taken and the outcome.

**Severity Scale for Adverse Events**:

|  |  |  |
| --- | --- | --- |
| **1** | **Mild** | Symptom(s) barely noticeable to the patient or causing no discomfort. The AE has no behavioral or functional effect. Treatment of the symptom(s) is generally not necessary. |
| **2** | **Moderate** | Symptom(s) of sufficient severity to inconvenience the patient. Daily activity is altered. Treatment of the symptoms may be necessary. |
| **3** | **Severe** | Symptom(s) of sufficient severity to seriously inconvenience the patient. The severity may lead to treatment discontinuation. Treatment of the symptoms may be necessary. |

**Relationship between Adverse Events and the Measurement Procedure**:

|  |  |  |
| --- | --- | --- |
| **0** | **Unrelated** | The chronological relationship between the occurrence of the clinical event and the performance of the measurement makes any causal link improbable, or other medications, therapeutic procedures, or underlying conditions provide a sufficient explanation for the observed event. |
| **1** | **Related** | The chronological relationship between the occurrence of the clinical event and the performance of the measurement makes a causal link possible. Furthermore, other medications, therapeutic procedures, or underlying conditions do not provide a sufficient explanation for the observed event. |

**6.2.1. Procedures to Follow in Case of a Serious Adverse Event**

**Investigator's Role(s)**

To ensure patient safety, any serious adverse event, whether or not the investigator suspects it to be related to the research or the study product, must be reported using the serious adverse event reporting form and sent by email to the Institut Régional de Médecine Physique et de Réadaptation de Nancy, an establishment of UGECAM du Nord-Est, without delay from the day the investigator becomes aware of it. The "Serious Adverse Event Reporting Form" found in the case report form must be completed, signed, and sent, possibly electronically, to the coordinating investigator, Christian Beyaert, and to the sponsor's representative, Jonathan Pierret. The investigator follows up on participants who have experienced a serious adverse event until the event is considered resolved.

**Sponsor's Role**

The Institut Régional de Réadaptation de Nancy will :

- Review the serious adverse event forms submitted by the investigator.
- Assess the possible relationship with the study product or research and its expectedness.
- Request additional information from the investigator if necessary.
- Complete the section reserved for the sponsor on the serious adverse event forms.
- In accordance with Article R1123-59 of the Public Health Code, the sponsor will promptly inform the competent authority and the Committee for the Protection of Persons of any new findings and, if applicable, of the measures taken.

**7. Data Management**

**7.1. Right of Access to Data**

The investigator agrees to allow free access to study data to the sponsor, monitors, auditors, or representatives of Health Authorities. At the end of the study, if the volunteer wishes, they may be informed of the overall results of the research. The participant must make a written request to the coordinator.

**7.2. Source Data**

Source documents are defined as any original document or object that can prove the existence or accuracy of a piece of data or a fact recorded during the clinical trial. They include original documents, data, and records from which the study data concerning the subject are transcribed into the case report form. This includes, among other things, the medical record, examination reports and results, questionnaires, and medical correspondence.

**7.3. Participant Anonymity**

The anonymity of participants will be ensured by assigning them a unique alphanumeric reference, for example, FLA-001 for the first participant. This reference will be used to name the files containing the data to be analyzed and to perform calculations and statistical studies. Video and photo recordings may be taken, subject to the participant's consent. The participant's anonymity will be ensured by masking their face and any distinguishing features.

**7.4. Confidentiality**

In accordance with the provisions of Article R. 5120 of the Public Health Code, the investigator and any person called upon to collaborate in the trials are bound by professional secrecy.

**7.5. Archiving**

At the end of the study, all study-related documents (including a copy of the case report forms) will be archived at the study sites or in a centralized archive. All study-related documents must be kept for 15 years after the end of the study.

**7.6. Rules for Publication of the Final Report**

This study will lead to publications and the writing of master's theses and/or dissertations. A summary of this final report will be sent to the CPP within 12 months following the end of the study.

**8. Quality Assurance**

**8.1. Investigator's Commitment - Good Clinical Practices**

The investigator undertakes to respect the protocol and to apply it taking into account good clinical practices. The investigator agrees to allow free access to study data to the sponsor, monitors, auditors, or representatives of Health Authorities.

**8.2. Data Quality Control**

Once the data are entered, data checks (consistency and missing data checks) will be carried out regularly for the clinical data. Missing or inconsistent data will be sent back to the investigators for additional information and verification.

**8.3. Audit and Inspection**

The investigators and the sponsor agree to comply with the requirements of these assessments, should they be carried out.

**9. Study Organization**

**9.1. Scientific Committee**

The scientific committee is led by Dr. Beyaert. Its objectives are to define the study's objective, write the protocol and the case report form, and select the centers. It will also ensure the smooth running of the study from a scientific and logistical standpoint, resolve any difficulties encountered, and make all decisions concerning the study (amendments, decisions on its premature termination or extension, re-evaluation of the study's sample size, etc.).

**10. Ethical and Regulatory Considerations**

**10.1. Research Site Authorization**

The IRR is a healthcare facility where the procedures performed in this research are those usually practiced as part of its activity.

**10.2. Ethical Conduct of the Study**

The planning and conduct of this study are governed by French and European laws (in particular, Decree No. 2016-1537 (Jardé Law) of November 16, 2016). The study will be conducted in accordance with the ethical principles of the Declaration of Helsinki and the recommendations of Good Clinical Practices.

**10.3. Committee for the Protection of Persons (CPP)**

The trial cannot begin without having received authorization from the CPP. The protocol will start upon receipt of a favorable opinion from the CPP. Any substantial modifications must be submitted as an amendment to the protocol and receive approval from the CPP before being implemented.

**10.4. Investigator's Responsibilities**

The investigator undertakes to ensure that this study is conducted in compliance with Law No. 2004-806 of August 9, 2004, on public health policy and its implementing decrees, the Declaration of Helsinki, and Good Clinical Practices. The investigator will inform volunteers about the study's objectives and constraints, and their right to refuse to participate or to withdraw at any time. The investigator will obtain written consent after ensuring the volunteer has understood the implications of participation.

**10.5. Sponsor's Responsibilities**

In accordance with the law, the sponsor commits to:

- **Insurance:** The sponsor has taken out insurance for the entire duration of the study to cover its own civil liability as well as that of any intervener involved in the trial.
- **Committee for the Protection of Persons:** The sponsor will seek the opinion of a CPP. The research can only be implemented after a favorable opinion from the CPP.
- **National Commission on Informatics and Liberty (CNIL):** This study falls under the MR-001 methodology. The Sponsor sent a declaration of conformity to the CNIL on 25/01/2021 under the number 2220869.

**11. Bibliography**

1.	Graham HK, Rosenbaum P, Paneth N, Dan B, Lin JP, Damiano DL, et al. Cerebral palsy. Nat Rev Dis Primers. 07 2016;2:15082.

2.	Pierret J, Beyaert C, Paysant J, Caudron S. How do children aged 6 to 11 stabilize themselves on an unstable sitting device? The progressive development of axial segment control. Human Movement Science. juin 2020;71:102624.

3.	Pin TW, Butler PB, Cheung HM, Shum SLF. Relationship between segmental trunk control and gross motor development in typically developing infants aged from 4 to 12 months: a pilot study. BMC Pediatr. déc 2019;19(1):425.

4.	Pin TW, Butler PB, Cheung HM, Shum SLF. Longitudinal Development of Segmental Trunk Control in Full Term and Preterm Infants- a Pilot Study: Part II. Developmental Neurorehabilitation. 2 avr 2020;23(3):193‑200.

5.	Saavedra SL, Woollacott MH. Segmental Contributions to Trunk Control in Children With Moderate-to-Severe Cerebral Palsy. Archives of Physical Medicine and Rehabilitation. juin 2015;96(6):1088‑97.

6.	Pierret J, Caudron S, Paysant J, Beyaert C. Impaired postural control of axial segments in children with cerebral palsy. Gait & Posture [Internet]. 8 mars 2021 [cité 9 mars 2021]; Disponible sur: https://www.sciencedirect.com/science/article/pii/S0966636221000977

7.	Heyrman L, Feys H, Molenaers G, Jaspers E, Monari D, Nieuwenhuys A, et al. Altered trunk movements during gait in children with spastic diplegia: Compensatory or underlying trunk control deficit? Research in Developmental Disabilities. 1 sept 2014;35(9):2044‑52.

8.	Attias M, Bonnefoy-Mazure A, Lempereur M, Lascombes P, De Coulon G, Armand S. Trunk movements during gait in cerebral palsy. Clinical Biomechanics. janv 2015;30(1):28‑32.

9.	Wallard L, Dietrich G, Kerlirzin Y, Bredin J. Balance control in gait children with cerebral palsy. Gait & Posture. mai 2014;40(1):43‑7.

10.	Kim CJ, Son SM. Comparison of Spatiotemporal Gait Parameters between Children with Normal Development and Children with Diplegic Cerebral Palsy. J Phys Ther Sci. 2014;26(9):1317‑9.

11.	Hsue BJ, Miller F, Su FC. The dynamic balance of the children with cerebral palsy and typical developing during gait. Part II: Instantaneous velocity and acceleration of COM and COP and their relationship. Gait & Posture. avr 2009;29(3):471‑6.

12.	Saether R, Helbostad JL, Adde L, Brændvik S, Lydersen S, Vik T. Gait characteristics in children and adolescents with cerebral palsy assessed with a trunk-worn accelerometer. Research in Developmental Disabilities. juill 2014;35(7):1773‑81.

13.	Meyns P, Kerkum YL, Brehm MA, Becher JG, Buizer AI, Harlaar J. Ankle foot orthoses in cerebral palsy: effects of ankle stiffness on trunk kinematics, gait stability and energy cost of walking. European Journal of Paediatric Neurology. 2020;

14.	Armand S, Watelain E, Mercier M, Lensel G, Lepoutre FX. Identification and classification of toe-walkers based on ankle kinematics, using a data-mining method. Gait & posture. 2006;23(2):240‑8.

15.	Beyaert C, Pierret J, Vasa R, Paysant J, Caudron S. Toe walking in children with cerebral palsy: a possible functional role for the plantar flexors. Journal of Neurophysiology. oct 2020;124(4):1257‑69.

16.	Worthen-Chaudhari L, Bing J, Schmiedeler JP, Basso DM. A new look at an old problem: Defining weight acceptance in human walking. Gait & Posture. janv 2014;39(1):588‑92.

17.	Colborne GR, Wright FV, Naumann S. Feedback of triceps surae EMG in gait of children with cerebral palsy: a controlled study. Archives of physical medicine and rehabilitation. 1994;75(1):40‑5.

18.	Nielsen JB, Christensen MS, Farmer SF, Lorentzen J. Spastic movement disorder: should we forget hyperexcitable stretch reflexes and start talking about inappropriate prediction of sensory consequences of movement? Exp Brain Res [Internet]. 7 mai 2020 [cité 29 mai 2020]; Disponible sur: http://link.springer.com/10.1007/s00221-020-05792-0

19.	Willerslev-Olsen M, Andersen JB, Sinkjaer T, Nielsen JB. Sensory feedback to ankle plantar flexors is not exaggerated during gait in spastic hemiplegic children with cerebral palsy. J Neurophysiol. févr 2014;111(4):746‑54.

20.	Lorentzen J, Willerslev-Olsen M, Hüche Larsen H, Farmer SF, Nielsen JB. Maturation of feedforward toe walking motor program is impaired in children with cerebral palsy. Brain. 1 mars 2019;142(3):526‑41.

21.	Perry J, Burnfield JM. Gait analysis: normal and pathological function. 2nd. Thorofare, NJ: Slack Incorporated. 2010;

22.	Correa TA, Schache AG, Graham HK, Baker R, Thomason P, Pandy MG. Potential of lower-limb muscles to accelerate the body during cerebral palsy gait. Gait & posture. 2012;36(2):194‑200.

23.	Kurz MJ, Stuberg WA, DeJong SL. Mechanical work performed by the legs of children with spastic diplegic cerebral palsy. Gait & Posture. mars 2010;31(3):347‑50.

24.	Pierret J, Beyaert C, Vasa R, Rumilly E, Paysant J, Caudron S. Rehabilitation of Postural Control and Gait in Children with Cerebral Palsy: the Beneficial Effects of Trunk-Focused Postural Activities. Developmental Neurorehabilitation. 23 mars 2023;1‑13.

25.	Neptune RR, Kautz SA, Zajac FE. Contributions of the individual ankle plantar flexors to support, forward progression and swing initiation during walking. Journal of Biomechanics. nov 2001;34(11):1387‑98.

26.	Heyrman L, Molenaers G, Desloovere K, Verheyden G, De Cat J, Monbaliu E, et al. A clinical tool to measure trunk control in children with cerebral palsy: The Trunk Control Measurement Scale. Research in Developmental Disabilities. nov 2011;32(6):2624‑35.

27.	Arı G, Günel MK. A Randomised Controlled Study to Investigate Effects of Bobath Based Trunk Control Training on Motor Function of Children with Spastic Bilateral Cerebral Palsy. International Journal of Clinical Medicine. 2017;08(04):205.

28.	Butler PB. A preliminary report on the effectiveness of trunk targeting in achieving independent sitting balance in children with cerebral palsy. Clinical Rehabilitation. 1998;12(4):281‑93.

29.	El Shemy SA. Trunk endurance and gait changes after core stability training in children with hemiplegic cerebral palsy: A randomized controlled trial. BMR. 28 nov 2018;31(6):1159‑67.

30.	ElBasatiny H, Abdelaziem A. Effect of Trunk Exercises on Trunk control, Balance and Mobility Function in Children with Hemiparetic Cerebral Palsy. IJTRR. 2015;4(5):236.

31.	Numanoğlu Akbaş A, Kerem Günel M. Effects of Trunk Training on Trunk, Upper and Lower Limb Motor Functions in Children with Spastic Cerebral Palsy: A Stratified Randomized Controlled Trial. Konuralp Tıp Dergisi. 28 juin 2019;253‑9.

32.	van Tittelboom V, Heyrman L, De Cat J, Algoet P, Peeters N, Alemdaroğlu-Gürbüz I, et al. Intensive Therapy of the Lower Limbs and the Trunk in Children with Bilateral Spastic Cerebral Palsy: Comparing a Qualitative Functional and a Functional Approach. JCM. 15 juin 2023;12(12):4078.

33.	Martín-Valero R, Vega-Ballón J, Perez-Cabezas V. Benefits of hippotherapy in children with cerebral palsy: A narrative review. European Journal of Paediatric Neurology. nov 2018;22(6):1150‑60.

34.	Moraes AG, Copetti F, Angelo VR, Chiavoloni LL, David AC. The effects of hippotherapy on postural balance and functional ability in children with cerebral palsy. Journal of Physical Therapy Science. 2016;28(8):2220‑6.

35.	Mutoh T, Mutoh T, Tsubone H, Takada M, Doumura M, Ihara M, et al. Impact of Long-Term Hippotherapy on the Walking Ability of Children With Cerebral Palsy and Quality of Life of Their Caregivers. Front Neurol. 13 août 2019;10:834.

36.	Santos de Assis G, Schlichting T, Rodrigues Mateus B, Gomes Lemos A, dos Santos AN. Physical therapy with hippotherapy compared to physical therapy alone in children with cerebral palsy: systematic review and meta‐analysis. Develop Med Child Neuro. févr 2022;64(2):156‑61.

37.	Palisano R, Rosenbaum P, Walter S, Russell D, Wood E, Galuppi B. Development and reliability of a system to classify gross motor function in children with cerebral palsy. Developmental Medicine & Child Neurology. 29 sept 2008;39(4):214‑23.

38.	Gouelle A, Mégrot F, Presedo A, Husson I, Yelnik A, Penneçot GF. The Gait Variability Index: A new way to quantify fluctuation magnitude of spatiotemporal parameters during gait. Gait & Posture. juill 2013;38(3):461‑5.

39.	Gouelle A, Rennie L, Clark DJ, Mégrot F, Balasubramanian CK. Addressing limitations of the Gait Variability Index to enhance its applicability: The enhanced GVI (EGVI). Tan MP, éditeur. PLoS ONE. 1 juin 2018;13(6):e0198267.

40.	McCoy SW, Bartlett DJ, Yocum A, Jeffries L, Fiss AL, Chiarello L, et al. Development and validity of the early clinical assessment of balance for young children with cerebral palsy. Developmental Neurorehabilitation. déc 2014;17(6):375‑83.

41.	Avery LM, Russell DJ, Rosenbaum PL. Criterion validity of the GMFM-66 item set and the GMFM-66 basal and ceiling approaches for estimating GMFM-66 scores. Dev Med Child Neurol. juin 2013;55(6):534‑8.

42.	Ma Y, Aslam R, Jester A. Development and validation of a WHO ICF compliant hand and upper limb assessment tool for children: The Reach Out questionnaire. Journal of Hand Therapy. août 2023;S0894113023000340.

43.	Yam WKL, Leung MSM. Interrater Reliability of Modified Ashworth Scale and Modified Tardieu Scale in Children With Spastic Cerebral Palsy. J Child Neurol. déc 2006;21(12):1031‑5.

44.	Beyaert C, Vasa R, Frykberg GE. Gait post-stroke: Pathophysiology and rehabilitation strategies. Neurophysiologie Clinique/Clinical Neurophysiology. nov 2015;45(4‑5):335‑55.

45.	Russell DJ, Avery LM, Rosenbaum PL, Raina PS, Walter SD, Palisano RJ. Improved Scaling of the Gross Motor Function Measure for Children With Cerebral Palsy: Evidence of Reliability and Validity. Physical Therapy. 1 sept 2000;80(9):873‑85.

46.	Russell DJ, Avery LM, Walter SD, Hanna SE, Bartlett DJ, Rosenbaum PL, et al. Development and validation of item sets to improve efficiency of administration of the 66-item Gross Motor Function Measure in children with cerebral palsy: Validation of GMFM-66 Item Sets. Developmental Medicine & Child Neurology. févr 2010;52(2):e48‑54.

47.	LaForme Fiss A, McCoy SW, Bartlett D, Avery L, Hanna SE, On Track Study Team. Developmental Trajectories for the Early Clinical Assessment of Balance by Gross Motor Function Classification System Level for Children With Cerebral Palsy. Physical Therapy. 1 févr 2019;99(2):217‑28.

48.	Joanna M, Magdalena S, Katarzyna BM, Daniel S, Ewa LD. The Utility of Gait Deviation Index (GDI) and Gait Variability Index (GVI) in Detecting Gait Changes in Spastic Hemiplegic Cerebral Palsy Children Using Ankle–Foot Orthoses (AFO). Children. 25 sept 2020;7(10):149.

49.	Prosser LA, Atkinson HL, Alfano JM, Leff M, Kessler SK, Gouelle A, et al. Normalizing step-to-step variability to age in children and adolescents with hemiplegia. Gait & Posture. oct 2022;98:6‑8.

50.	Kurz MJ, Arpin DJ, Corr B. Differences in the dynamic gait stability of children with cerebral palsy and typically developing children. Gait & Posture. juill 2012;36(3):600‑4.

51.	Mancinelli C, Patel S, Deming LC, Schmid M, Patritti BL, Chu JJ, et al. Assessing the feasibility of classifying toe-walking severity in children with cerebral palsy using a sensorized shoe. In: 2009 Annual International Conference of the IEEE Engineering in Medicine and Biology Society [Internet]. Minneapolis, MN: IEEE; 2009 [cité 13 sept 2023]. p. 5163‑6. Disponible sur: http://ieeexplore.ieee.org/document/5332733/

**12. Annexes**

**12.1. Annex 1: List of Investigators and Scientific Collaborators**

**Investigators**

| First Name, Last Name | Title/Position | RPPS Number | Address |
| --- | --- | --- | --- |
| Christian, Beyaert | PU-PH, Pediatrician, Coordinating Investigator | 10002353976 | Center for Physical Medicine and Rehabilitation for Children, IRR of Nancy, 46 rue du Doyen Parisot, 54630 Flavigny-sur-Moselle |
| Fanny Dalmont | PRM Physician, Physician Investigator | 10101432648 | Center for Physical Medicine and Rehabilitation for Children, IRR of Nancy, 46 rue du Doyen Parisot, 54630 Flavigny-sur-Moselle |
| Marion, Birck | PRM Physician, Investigator | 10101658424 | Center for Physical Medicine and Rehabilitation for Children, IRR of Nancy, 46 rue du Doyen Parisot, 54630 Flavigny-sur-Moselle |

**Scientific Collaborators**

| First Name, Last Name | Title/Position | Address |
| --- | --- | --- |
| Jonathan, Pierret | PhD, Head of IRR Research Unit | Regional Institute of Physical Medicine and Rehabilitation of Nancy, 75 Boulevard Lobau, 54042, NANCY CEDEX |
| Sandrine, Regef | PRM Physician, CMPRE Coordinating Physician | Center for Physical Medicine and Rehabilitation for Children, IRR of Nancy, 46 rue du Doyen Parisot, 54630 Flavigny-sur-Moselle |
| Christelle Requena | Clinical Research Assistant – UGECAM du Nord-Est | Regional Institute of Physical Medicine and Rehabilitation of Nancy, 75 Boulevard Lobau, 54042, NANCY CEDEX |
| Stella Zografou | Physical Therapist | Regional Institute of Physical Medicine and Rehabilitation of Nancy, 75 Boulevard Lobau, 54042, NANCY CEDEX |

**12.2. Annex 2: List of Clinical Investigation Sites**

CMPRE of Flavigny-Sur-Moselle, Regional Institute of Physical Medicine and Rehabilitation (IRR), an establishment of UGECAM du Nord-Est, 46 Rue du Doyen Jacques Parisot BP2, 54630 Flavigny-sur-Moselle.
