## Supplementary material for "Investigating rehabilitation by activities involving the trunk to improve balance and gait control in young children with cerebral palsy: a randomized open-label crossover trial protocol": RAIT in CP CPP ENGLISH

**Ethics Committee Approval**

**File Reference:** 24.00651.000276 – 2023-A01969-36 Page 1 of 4

**East I Ethics Committee for the Protection of Persons**

**Opinion on Initial Application**

**CPP**

- **Committee Name:** East I Ethics Committee for the Protection of Persons
- **Address:** La Chartreuse Hospital Center - 1 Bld Chanoine Kir, BP 23314 21033 DIJON France

**Sponsor / Applicant**

- **Sponsor:** Regional Institute of Physical Medicine and Rehabilitation of Nancy - UGECAM North East Establishment
- **Legal Representative (EU):** -
- **Mandatary:** -

**File Details**

- **SI Number:** 24.00651.000276
- **National Number:** 2023-A01969-36
- **Internal Reference:** IRR-Flavigny-2023-2
- **Regulation:** Jardé Law
- **Classification:** Category 2
- **Product or Procedure:** Outside health products (products not mentioned in article L.5311-11 of the public health code)
- **Principal Investigator:** Christian Beyaert
- **Title:** Investigating rehabilitation by activities involving the trunk in young children with cerebral palsy

This file was reviewed in session on 11/04/2024 and the committee president was mandated to issue the opinion upon receipt of the applicant's responses to the latest requests. In view of the responses obtained, the following opinion has been issued. This opinion runs from the status change on the SI.

Considering that the ethical conditions are met, particularly with regard to the elements of article L.1123-7 of the public health code, the committee's examination allows to conclude that the research can be carried out and to render the following opinion:

**FAVORABLE OPINION**

This opinion is valid for two years. In accordance with article L.1123-11 of the public health code, the sponsor must declare the start of research to the CPP. This declaration is made directly on SIRIPH2G (using the "start study" button).

If you have not been able to include a first research participant within this timeframe, you may request an extension of this opinion from the CPP before its validity expires (article R.1123-26 of the public health code).

**Committee Members Who Deliberated**

| **College** | **Category** | **Name** | **Function** |
| --- | --- | --- | --- |
| College I | RIPH Qualification - Other | QUENOT Jean-Pierre | President |
| College II | Approved Association Representative | LECOMTE Yann | Vice-President |
| College I | RIPH Qualification - Biostatistics or Epidemiology | WHITE Olivier |  |
| College I | RIPH Qualification - Other | CHAVANET Pascal |  |
| College I | RIPH Qualification - Other | LAROCHE Davy |  |
| College I | RIPH Qualification - Other | GUILLOTEAU Adrien |  |
| College I | General Medicine Specialist | DURAND Rémy |  |
| College II | Ethics Competence | BOGGIO Vincent |  |
| College II | Legal Competence | TABUTIAUX Agnès |  |
| College II | Approved Association Representative | LEGENDRE Christiane |  |
| College II | Approved Association Representative | PLASSARD Françoise |  |

**Documents Analyzed by the CPP**

| **Category** | **Title** | **Submission Date** |
| --- | --- | --- |
| ADD - Additional Doc | 2023-A01969-36_Additional_v1_20240208.pdf | 13/02/2024 |
| ASS - Insurance | 2023-A01969-36_Insurance_v1_20240212.pdf | 13/02/2024 |
| ATT - Certificate | 2023-A01969-36_Receipt_MR001_CNIL_v1_20240208.pdf | 13/02/2024 |
| COU - Letter | 2023-A01969-36_Letter_v1_20240208.pdf | 13/02/2024 |
| CVI - Investigator CVs | 2023-A01969-36_Investigator_List_CV_v2_20240216.pdf | 16/02/2024 |
| DEM - Authorization Request | 2023-A01969-36_Request_v2_20240216.pdf | 16/02/2024 |
| DOC - Other Documents | 2023-A01969-36_Home_Follow-up_Sheet_parents-physio_v1_20240325.pdf | 27/03/2024 |
| DOC - Other Documents | 2023-A01969-36_Info_on_HR_Physio_v1_20240325.pdf | 27/03/2024 |
| INF - Information Doc | 2023-A01969-36_NIFC-Parents-TD_v2_20240216.pdf | 16/02/2024 |
| INF - Information Doc | 2023-A01969-36_NIFC-ParentsCP_v2_20240216.pdf | 16/02/2024 |
| INF - Information Doc | 2023-A01969-36_NIFC_physios_v1_20240325.pdf | 27/03/2024 |
| INF - Information Doc | 2023-A01969-36_NIFC-Parents-TD_v3_20240325-final.pdf | 27/03/2024 |
| INF - Information Doc | 2023-A01969-36_NIFC-Parents-TD_v3_20240325-follow-up.pdf | 27/03/2024 |
| INF - Information Doc | 2023-A01969-36_NIFC-ParentsCP_v3_20240325-final.pdf | 27/03/2024 |
| INF - Information Doc | 2023-A01969-36_NIFC-ParentsCP_v3_20240325-follow-up.pdf | 27/03/2024 |
| INF - Information Doc | 2023-A01969-36_NIFC_physios_v2_20240418-final.pdf | 18/04/2024 |
| INF - Information Doc | 2023-A01969-36_NIFC_physios_v2_20240418-follow-up.pdf | 18/04/2024 |
| INF - Information Doc | 2023-A01969-36_NIFC-Parents-TD_v4_20240418-final.pdf | 18/04/2024 |
| INF - Information Doc | 2023-A01969-36_NIFC-Parents-TD_v4_20240418-follow-up.pdf | 18/04/2024 |
| INF - Information Doc | 2023-A01969-36_NIFC-ParentsCP_v4_20240418-final.pdf | 18/04/2024 |
| INF - Information Doc | 2023-A01969-36_NIFC-ParentsCP_v4_20240418-follow-up.pdf | 18/04/2024 |
| JUS - Research Site Justification | 2023-A01969-36_Equipment_v1_20240802.pdf | 13/02/2024 |
| LET - Letter | 2023-A01969-36_Recruitment-Email_TD_v1_20240325.pdf | 27/03/2024 |
| PRO - Protocol | 2023-A01969-36_Protocol_v1_20240208.pdf | 13/02/2024 |
| PRO - Protocol | 2023-A01969-36_Protocol_v2_20240325-follow-up.pdf | 27/03/2024 |
| PRO - Protocol | 2023-A01969-36_Protocol_v2_20240325-final.pdf | 27/03/2024 |
| PRO - Protocol | 2023-A01969-36_Protocol_v3_20240418-final.pdf | 18/04/2024 |
| PRO - Protocol | 2023-A01969-36_Protocol_v3_20240418-follow-up.pdf | 18/04/2024 |
| QUE - Scales/Questionnaires | 2023-A01969-36_Reach-Out-5-9years_v1_20240208.pdf | 13/02/2024 |
| QUE - Scales/Questionnaires | 2023-A01969-36_GMFM-66-SI-v1_20240208.pdf | 13/02/2024 |
| QUE - Scales/Questionnaires | 2023-A01969-36_ECAB-v1-20240208.pdf | 13/02/2024 |
| QUE - Scales/Questionnaires | 2023-A01969-36_Reach-Out-2-4years_v1-20240208.pdf | 13/02/2024 |
| QUE - Scales/Questionnaires | 2023-A01969-36_Reach_Out-2-4years_v2-20240325.pdf | 27/03/2024 |
| QUE - Scales/Questionnaires | 2023-A01969-36_Reach_Out-5-9years_v2_20240325.pdf | 27/03/2024 |
| REP - Response Letter | 2023-A01969-36-Response-Letter_v1_20240325.pdf | 27/03/2024 |
| REP - Response Letter | 2023-A01969-36-Response-Letter_v2_20240418.pdf | 18/04/2024 |
| RES - Summary | 2023-A01969-36_Summary_v1_20240208.pdf | 13/02/2024 |
| RES - Summary | 2023-A01969-36-Summary_v2_20240325-modified.pdf | 27/03/2024 |
| RES - Summary | 2023-A01969-36-Summary_v2_20240325-final.pdf | 27/03/2024 |

**Notes:**

- *Documents labeled as non-compliant on the RIPH2G SI or transmitted for information/notification as part of this opinion request were not evaluated by the CPP.
- *The titles of documents examined by the committee, listed in this opinion, use the file nomenclature used by the applicant on the RIPH2G SI.

**President of CPP EST 1**
**Prof. Jean-Pierre QUENOT**
